# Predicting Polycystic Ovary Syndrome among Reproductive-Aged Women in Bangladesh Using Machine Learning Algorithms: Development of a Hospital-Based Predictive Model

**DOI:** 10.1101/2025.11.19.25340618

**Authors:** Anup Talukder, Tahmina Akter Tithi, Abdul Muyeed, Md. Shahriar Hossain, Mahedi Hasan Nahid

## Abstract

**Background:** Polycystic ovary syndrome (PCOS) is one of the most common endocrine disorders among reproductive aged women, which is characterized by hormonal imbalance, metabolic disorders, and reproductive complications. Despite increasing rates in South Asia, notably Bangladesh, diagnostic constraints and fragmented data make early detection difficult. Machine learning (ML) may provide a solution by detecting subtle trends in clinical, demographic, and psychological data to improve diagnostic accuracy and timely intervention.

**Objective:** The study aimed to predict PCOS using machine-learning approaches among reproductive women of Bangladesh and to quantify the relative predictive contribution of psychological and clinical predictors to disease diagnosis.

**Methods:** A cross-sectional survey with 212 reproductive aged women was conducted to evaluate machine-learning models for predicting PCOS. Data were collected using the DASS-21 and ISI-7 scale and feature selection was performed using the LASSO regression. Model performance was evaluated by bootstrapping and nested cross-validation to ensure robustness. Several machine-learning models were developed, and predictive performance was assessed using various evaluation metrics.

**Results:** Nearly half of the participants (49.5%) were diagnosed with PCOS. Out of 21 features, eight were selected as significant by LASSO. Extreme Gradient Boosting (XGBoost) demonstrated best predictive performance with an accuracy of 99.63%, sensitivity of 99.45%, specificity of 99.81%, Cohen’s Kappa of 99.26%, and ROC-AUC of 99.99% among all models. Random Forest and Support Vector Machine also showed strong results, confirming the effectiveness of ensemble and kernel-based approaches. Moreover, psychological aspects showed a minimal predictive influence than clinical features.

**Conclusions:** Using ML frameworks to incorporate clinical, demographic, and psychological data can significantly enhance PCOS prediction in contexts with limited resources. The XGBoost model demonstrated remarkable reliability and accuracy, underscoring its efficacy as a tool for clinical decision support. Future studies should include biochemical indicators for wider application in the management of women’s reproductive health and globally validate these findings.

## 1.0 Introduction

Polycystic ovary syndrome (PCOS) is a complex endocrine disease that impacts women in reproductive age. The signs of PCOS are ovarian dysfunction, irregular menstruation and infertility. It may also cause hirsutism, acne, androgenetic alopecia and small ovarian cysts [1,2]. It may make the health of women complicate by causing hormonal abnormalities. High insulin levels and Insulin resistance are significant issues and risk factors [3]. There are several causes that include genetic factors. It tends to appear in families and certain gene variations in the CYP11A1, FSHR, LHCGR, and INSR systems seem to be involved [4]. Lifestyle factors like obesity, poor diet and not enough exercise play a key role [1]. PCOS is related to many physical problems like metabolic syndrome, type 2 diabetes, raised cholesterol, and a greater chance of heart issues [5]. External symptoms and complications can cause psychological distress including anxiety, depression and lack of self-confidence. This can result in social stigma and a lower quality of life [6]. PCOS has a large economic burden. It is related to the direct and indirect costs because of health expenditure, lost productivity, and life-long management of individuals with the condition. It is costing already billions of dollars annually and it is clearly a major social health problem [7].

Rotterdam (2003) criteria, NIH (1990) and AE PCOS guidelines (2006) are all framework materials utilized for PCOS diagnosis [8]. The 2023 guideline based on international evidence clarified the Rotterdam criteria’s diagnostic approach. It requires the presence of two from hyperandrogenism, ovulatory dysfunction and the typical polycystic ovarian appearance [9]. Globally, the prevalence is between 6% and 21%. It differs significantly in recent systemic reviews and also depends on the diagnostic criteria used, demographics of the screened population and techniques employed in the studies [10]. This broad range of prevalence highlights the complexity of PCOS. However, this burden of PCOS is not equally distributed globally. It shows a difference between developed (Global North) and developing (Global South) countries. In a key 2022 meta-analysis of developed nation populations, specifically in Europe and USA, the prevalence estimates ranged widely by diagnostic lens. NIH criteria showed a prevalence of 6.2, whereas the prevalence according to Rotterdam criteria was 19.5% [11]. A recent study has shown that having PCOS increases depression and anxiety risk by 65% and 42% respectively with an estimated annual cost of 4.26 billion dollars [12]. Since these developed countries provide a high level of diagnostic facilities such as hormonal tests, imaging and they also possess strong level of electronic health record, which enables reliable data to be gathered in research and clinical studies [13,14]. Previous studies have found that prevalence has continued to rise in the Middle Eastern and North African countries between 1990 and 2019. In 2019, region’s age-standardized prevalence was 2,079.7 per 100,000 populations, representing a 37.9% increase over the period [15]. A study found women with PCOS had more chances of experiencing unhealthy mental and sleep habits, poor dietary practices and PMS symptoms in Palestine [16]. Global data has indicated that the prevalence is rising in socioeconomically developed locations. The primary causes for this trend are some factors including Lifestyle changes, dieting and lack of physical exercise [13].

In particular, the increasing prevalence of PCOS related to modern lifestyle is becoming a significant health issue in Asia where South Asia has emerged as one of the global hotspots in PCOS [17]. In 2024, from a study of 20 studies and 23 countries including 14,010 adolescent participants found that approximately 11.4% of women in Southeast Asia have PCOS [17]. A Previous study showed that the prevalence in south Asia has increased by 1.87% in a year [18]. Researchers have shown that according to the Rotterdam criteria, 17.4% of women in Delhi, 8.1% of women in Chennai and 22.5% of women in Mumbai had PCOS which indicates the significant impact of local lifestyles and environmental factors [19–21].In an Indian study, Depression (66.1%) and anxiety (56.9%) were significantly more prevalent in PCOS than the control [22]. Using the Rotterdam criteria, a hospital-based study conducted among infertile women in Nepal showed a PCOS prevalence of 7.3% [23]. In Pakistan, A previous study showed the prevalence in adolescent girls of 13-19 years is 26.7% based on Rotterdam criteria. Moreover, urban-rural differences were highlighted in the investigation with a range of 34.1 percent of urban areas and 19.0 percent of rural areas [24]. A study in Pakistan has also shown that high rates of anxiety and depression with social and economic factors are also important [25].

Among South Asian regions, Bangladesh has some tough challenges where the high prevalence of PCOS necessitates particular attention to research. Recent Bangladeshi research has found that the prevalence of PCOS ranging from 6.1%-92.2% in clinic populations, 29.9%-46.2% among infertile women and 37% in medical students [26]. The most research of Bangladesh on PCOS is mainly hospital-based. A study has shown high rates of obesity (69%), dyslipidemia (91%) and metabolic syndrome (42%) in an adolescent cohort [27]. The prevalence of PCOS among Bangabandhu Sheikh Mujib Medical University (BSMMU) outpatients was 6.1% and it was 35.4% among infertile women [28]. Another BSMMU study found dyslipidemia in 93.7% and metabolic syndrome in 42.9% [29]. A study from BSMMU reports that 68% experience anxiety, 32% depression and 45% stress [6]. On a countrywide online survey, 71% reported loneliness, 88% anxiety and 60% depression [30]. PCOS also creates widespread stigma in society where symptoms such as infertility, irregular menstruation, hirsutism and weight gain are viewed negatively in society. This increased social pressure discourages early medical interactions and limits socialization of the affected women [31]. In Mymensingh, A sonographic study has found 12.5% prevalence of symptomatic women. Its incidence was more among women in the reproductive age in Mymensingh area, Bangladesh of 21-25 [32]. The wide range of prevalence rates defines the difficulty of incomplete data, absence of large-scale and community-based epidemiological research in Bangladesh.

Previous investigations reveal that PCOS is a global concern, but there is a significant gap between research and diagnostic opportunities in developed and developing countries. At the same time, developed countries use advanced diagnostic methods and robust electronic health records to control PCOS [13,14]. These disparities are particularly evident in Bangladesh, which faces challenges including fragmentation of data, over dependence on hospital-based studies and absence of large-scale population based epidemiological research [26]. This lack of data not only obscures the true picture of PCOS but also prevents timely and accurate diagnosis, as older methods do not assess the extent of the problem. Machine learning (ML) is globally used to identify PCOS at an early phase through detecting patterns in clinical and lifestyle data [14]. Moreover, the study has the potential to support the WHO/UN’s third Sustainable Development Goal of good health and well-being. This can be done through improvement in reproductive health outcomes and fewer cases of non-communicable diseases [33,34]. Models that we developed in our study can identify PCOS quickly, accurately and ultimately help initiate timely treatment. Such innovations are essential to reduce the chances of long-term health problems related to PCOS. As a result, this AI based methods save the health status of at-risk women and contributes greatly to enhancing the quality of life and promoting the overall health benefits of the population.

In order to fill this research gap, the study aims to develop machine-learning models to accurately detect and predict PCOS among reproductive aged women in Bangladesh. The specific objectives are to quantify the relative predictive contribution of clinical, demographic, and lifestyle factors of PCOS and to construct predictive models based on these findings. By addressing these objectives, the study seeks to find a possible solution for the problem of PCOS in Bangladesh different from standard approaches. It also aims to develop the skills of the doctors with new tools, which will improve health, and well-being of millions of women in Bangladesh.

## 2.0 Materials and Method

### 2.1. Ethics Statement

Ethical approval for this study was granted by the Research Ethics Committee of the Public Health Foundation Bangladesh (PHFBD) in Dhaka. The study adhered to the ethical principles outlined in the Declaration of Helsinki. Written informed consent was obtained from all participants prior to data collection. The research protocol was approved under reference number PHFBD-ERC-NFP-E-18/2025.

At the beginning of data collection, participants received a thorough explanation of the study’s objectives, aims, methods, affiliations, benefits, and hazards at the planned face to face interview session. Afterward, the respondents agreed to take part in the study, and the data collectors promised that their answers would be kept private and not shared. With their consent, data was subsequently gathered from them for the research purpose.

### 2.2. Study design

A cross-sectional study design was conducted to evaluate machine-learning models for predicting polycystic ovary syndrome (PCOS). Zad Z et al. (2024) and Elmannai H et al. (2023) also used comparable methodologies in their study for predicting PCOS [14,35].

### 2.3. Study participants and Sample

The study participants were the women of reproductive age. Regardless of whether they had PCOS or not, no respondents under the age of 18 were included in the final sample, despite the reason that the reproductive age range was set at 15–49 years [36]. The Godden’s formula was utilized to calculate the sample size considered a 5% margin of error and a 5% significance level (α = 0.05). It was determined using a cross-sectional survey conducted by Keeratibharat P et al. (2024) that reported an 11.92% prevalence of anxiety among Thai women with PCOS [37].

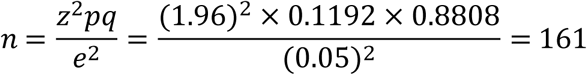

Using the formula, the optimal sample size was found 161. The sample size of this study is consistent with a previous clinical machine learning work using a small pediatric HIV cohort, which shows that ML models can be utilized reliably even with small sample size if adequate validation procedures are implemented [38]. This evidence justified the calculated sample size for generating exploratory predictive models aimed at identifying patterns related to PCOS. Initially, the questionnaire was given to 220 participants despite of knowing whether they had PCOS or not. After that, 212 participants were finally selected for the study with their consent after eight individuals were excluded due to missing data, and individuals with acute mental issues, hormonal medication, and metabolic anomalies. Among the 212 participants, 105 individuals had PCOS by satisfying at least two Rotterdam criteria. On the other hand, 107 participants did not have PCOS because they met less than two Rotterdam criteria. Figure 1 depicts the exclusion and inclusion criteria of sampling of the study.

**Fig 1.**
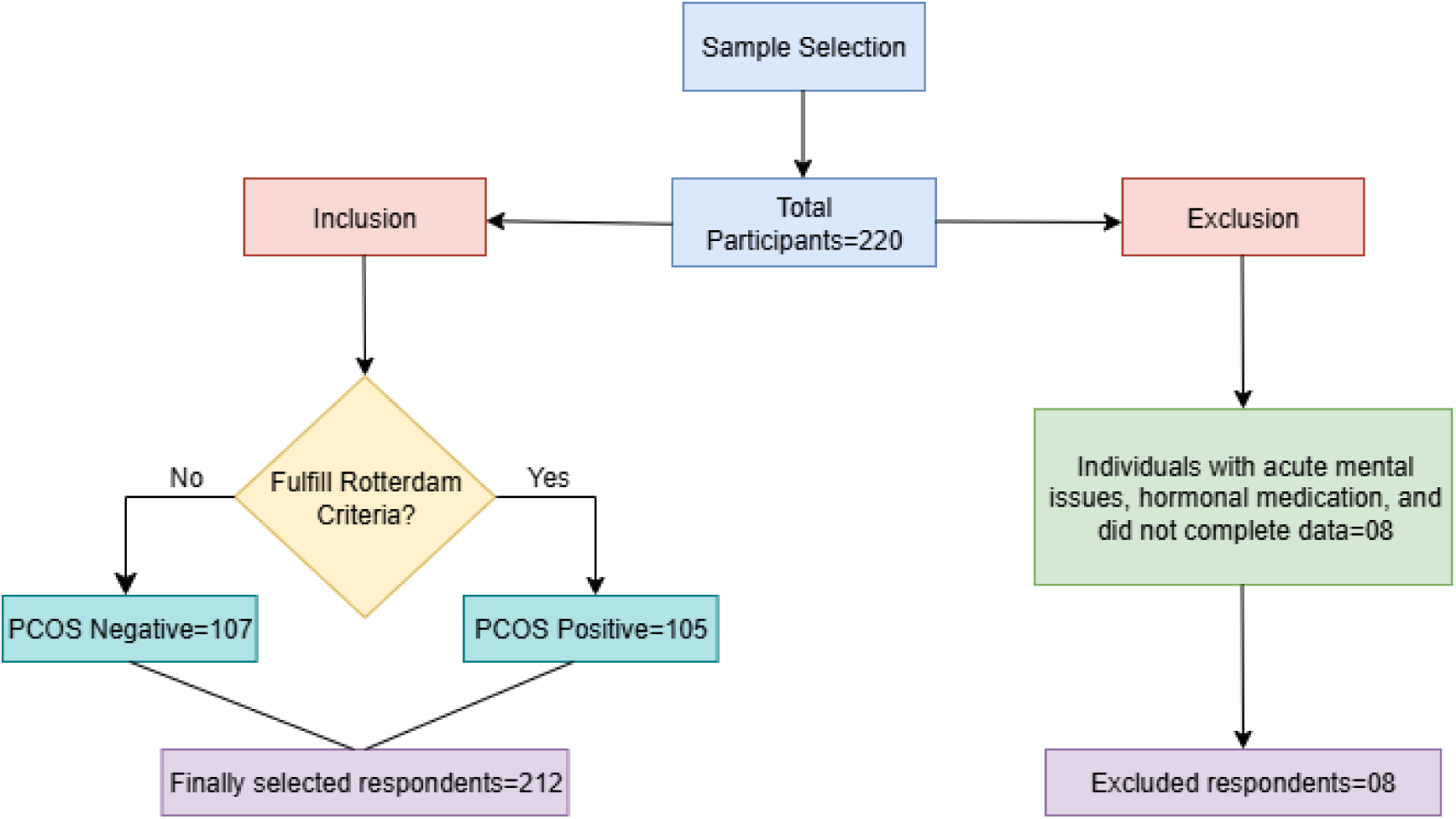
Sampling Flow chart with study inclusion and exclusion criteria.

A post-hoc power analysis was performed to determine if the appropriate sample size of 105 PCOS participants and 107 non-PCOS participants had enough power to identify group differences in psychological distress. The results showed an estimated power of around >95%, indicating that there was enough data in the sample to detect an odds ratio of 4.6. Notably, the power was significantly lower for detecting smaller effect sizes. Furthermore, the study was strongly powered, as evidenced by the substantial effect of 63% that was discovered when estimating the effect size using Cohen’s h. Consequently, optimal sample sizes provide enough power to assess moderate-to-large correlations.

### 2.4. Instrument and data collection

The study participants were selected from the outpatients of the gynecology departments of four government hospitals in the Mymensingh division using a convenience selection technique. The data was gathered between March 1, 2025, and April 15, 2025. As in Bangladesh, the state of reproductive health are almost identical in every administrative division [39], the present study was conducted on Mymensingh conveniently for reducing cost and time. The study’s area map was shown in Figure 2. Data were obtained from four government hospitals situated over four administrative districts in the Mymensingh division. These include Mymensingh Medical College and Hospital, Sherpur Sadar Hospital, Jamalpur Sadar Hospital and, Netrokona Medical College and Hospital. Before the data collection, we made the decision to recruit about similar numbers of participants (50 each) from each hospital in order to ensure equitable representation from all four districts of the Mymensingh division. As Mymensingh Medical College and Hospital is the largest referral centre with the most patients, a marginally greater number (62) of sample was collected from here. Due to logistical considerations such staff availability, time, and resources, as well as the need for fair representation across sites, this quota-based approach was employed before data collection. Data was collected from the reproductive-aged women who were leaving the gynecology departments and met the inclusion criteria after getting their consent. In order to gather data, an in-person interview was conducted, and the responses were recorded using a structured questionnaire. A four-section structured questionnaire in a printed version was given to the participants, which was developed in Bengali and English. To ensure the reliability and validity of the study, a pilot study was conducted on 10 participants. In the first section of the questionnaire, the Rotterdam criteria were included. Rotterdam uses three criteria to diagnose PCOS: polycystic ovaries on ultrasound, hyperandrogenism, and irregular menstruation. PCOS had to be diagnosed based on at least two of these three requirements. The diagnosis of PCOS was verified by the trained gynecologists at each of the four participating hospitals to maintain consistency. Clinicians attest that clinical criteria, specifically hirsutism, acne, and androgenic alopecia as stated by participants, were utilized to evaluate hyperandrogenism. Biochemical testing for androgen levels was not carried out due to a lack of resources. In accordance with the Rotterdam criteria, irregular menstruation was categorized as either persistent irregular periods (less than 8 cycles per year), amenorrhoea (absence of menstruation for more than 90 days), or oligomenorrhea (cycles longer than 35 days). Polycystic ovaries were confirmed by ultrasound when the ovarian volume exceeded 10 cm³ or each ovary contained ≥12 follicles measuring 2–9 mm.

**Fig 2.**
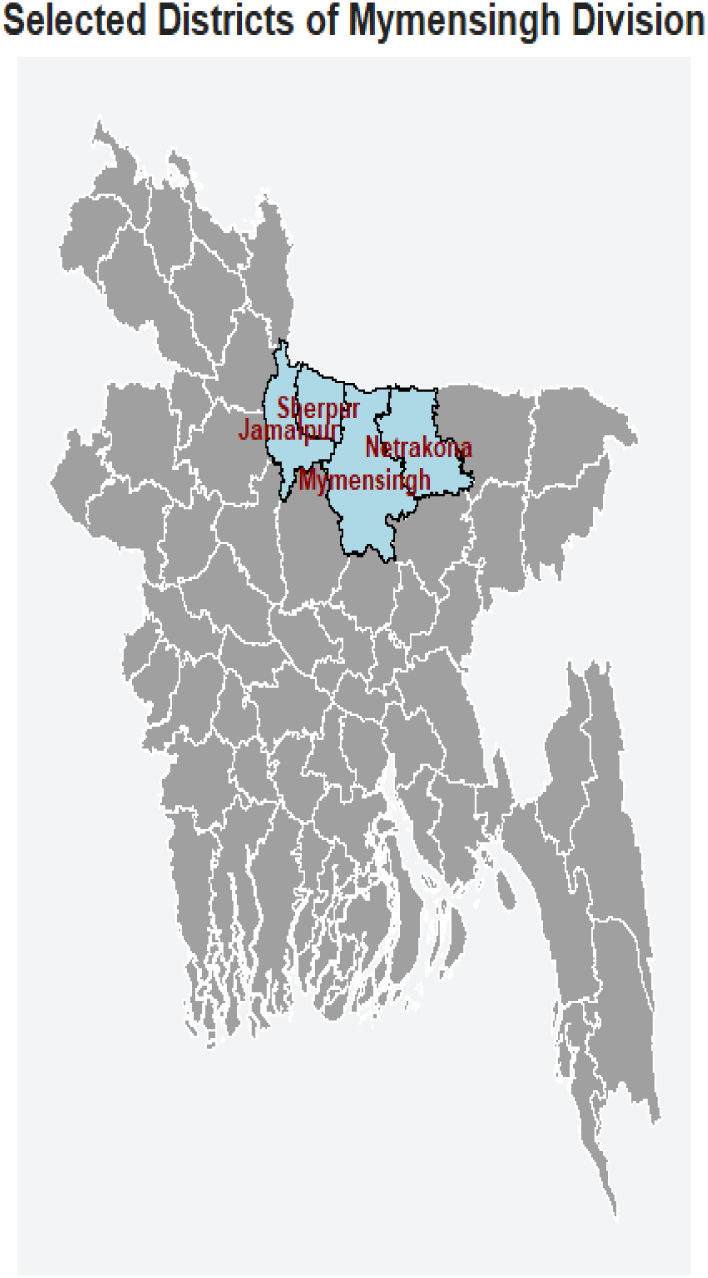
Map of the study area.

In the second, third and final section of the questionnaire included the background information, the depression, anxiety, and stress scale (DASS-21 scale), and the insomnia severity index (ISI-7) scale respectively.

### 2.5. Depression, Anxiety, Stress Scale-21 (DASS-21)

The Depression, Anxiety, and Stress Scale-21 is a self-reported measurement tool that are used to assess the emotional states of depression, anxiety, and stress of individuals over the past week. Each of the three scales has seven items, which are separated into subscales with comparable content. It contains 4 point likert scale ranging from 0 to 3 to rate the level of agreement. The number 0,1,2,3 represent not at all, some of the time, good part of the time, and most of the time respectively. The scores of the relevant items are summed and multiplied by two to get the final result [40]. Table_1 illustrates the severity labels for depression, anxiety, and stress after calculating the score. The severity label that ranges from moderate to extremely severe indicates hazardous scores for depression, anxiety, and stress. In the pilot study, this scale had a Cronbach’s α of 0.76, indicating a good reliability and internal consistency. The tool’s final version retained this level of reliability after data collection. As part of the language translation process, a faculty member from Jatiya Kabi Kazi Nazrul Islam University’s English department verified the Bengali translation of the original DASS-21 scale to make sure it was equivalent.

**Table 1.**
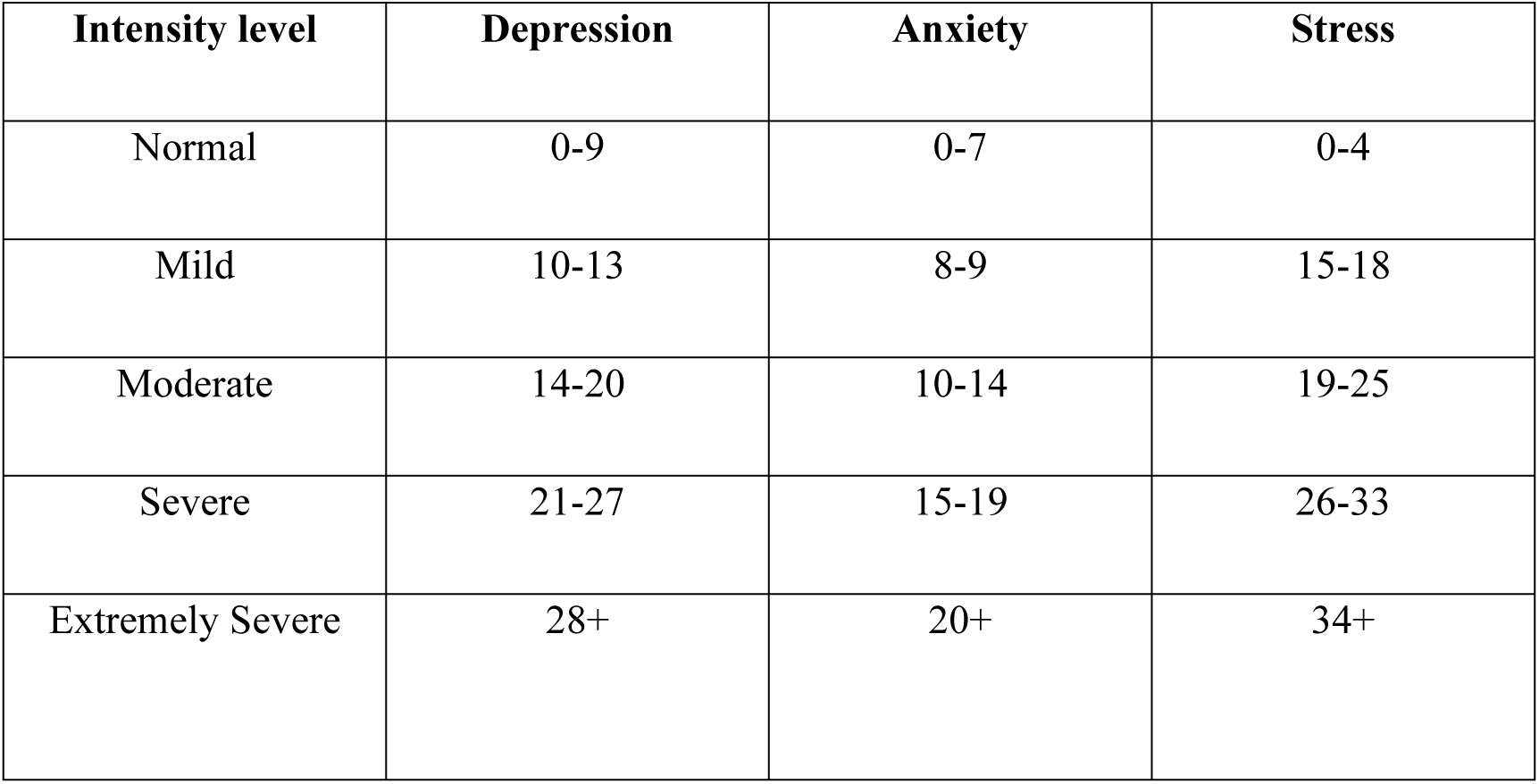
Cut-off scores of DASS-21.

| Intensity level | Depression | Anxiety | Stress |
| --- | --- | --- | --- |
| Normal | 0-9 | 0-7 | 0-4 |
| Mild | 10-13 | 8-9 | 15-18 |
| Moderate | 14-20 | 10-14 | 19-25 |
| Severe | 21-27 | 15-19 | 26-33 |
| Extremely Severe | 28+ | 20+ | 34+ |

### 2.6. Insomnia Severity Index (ISI-7)

The Insomnia Severity Index (ISI-7) is a widely used self-reported screening tool to assess the level of insomnia of an individual over the last two weeks. It consists of seven items questionnaire to rate one’s sleep problem using a five-point likert scale. All the seven items scores are summed to get the final score, where total score ranges from 0 to 28. Higher scores indicate more severe problem of insomnia. The score ranges from 0 to 7, indicating no clinically significant insomnia. Similarly, the score ranges from 8 to 14, 15 to 21, and 22 to 28, indicating subthreshold insomnia, clinical insomnia (moderate severity), and clinical insomnia (severe) respectively [41]. The moderate and severe levels of insomnia are used to determine the problematic scores. A professor from the English department at Jatiya Kabi Kazi Nazrul Islam University verified the original ISI scale’s Bengali translation as part of the validation process. In the pilot test, the scale’s Cronbach’s α coefficient was 0.888, demonstrating high internal consistency and reliability. This level of reliability was maintained in the tool’s final version after data collection.

### 2.7. Statistical Analysis

The study used DASS-21 and ISI-7 to measure the psychological distress and insomnia level respectively. Cronbach’s 𝛼 was used to measure the internal consistency for assessing the reliability of the tools. Moreover, descriptive statistics was used to summarize the socio-demographic, PCOS, and psychological factors related characteristics. The workflow of the study is presented in Figure 3. The primary workflow followed pre-processing, feature selection, model fitting, simulate data, and model evaluation. All statistical analyses were implemented using R (version 4.5.1) software, and Python.

**Fig 3.**
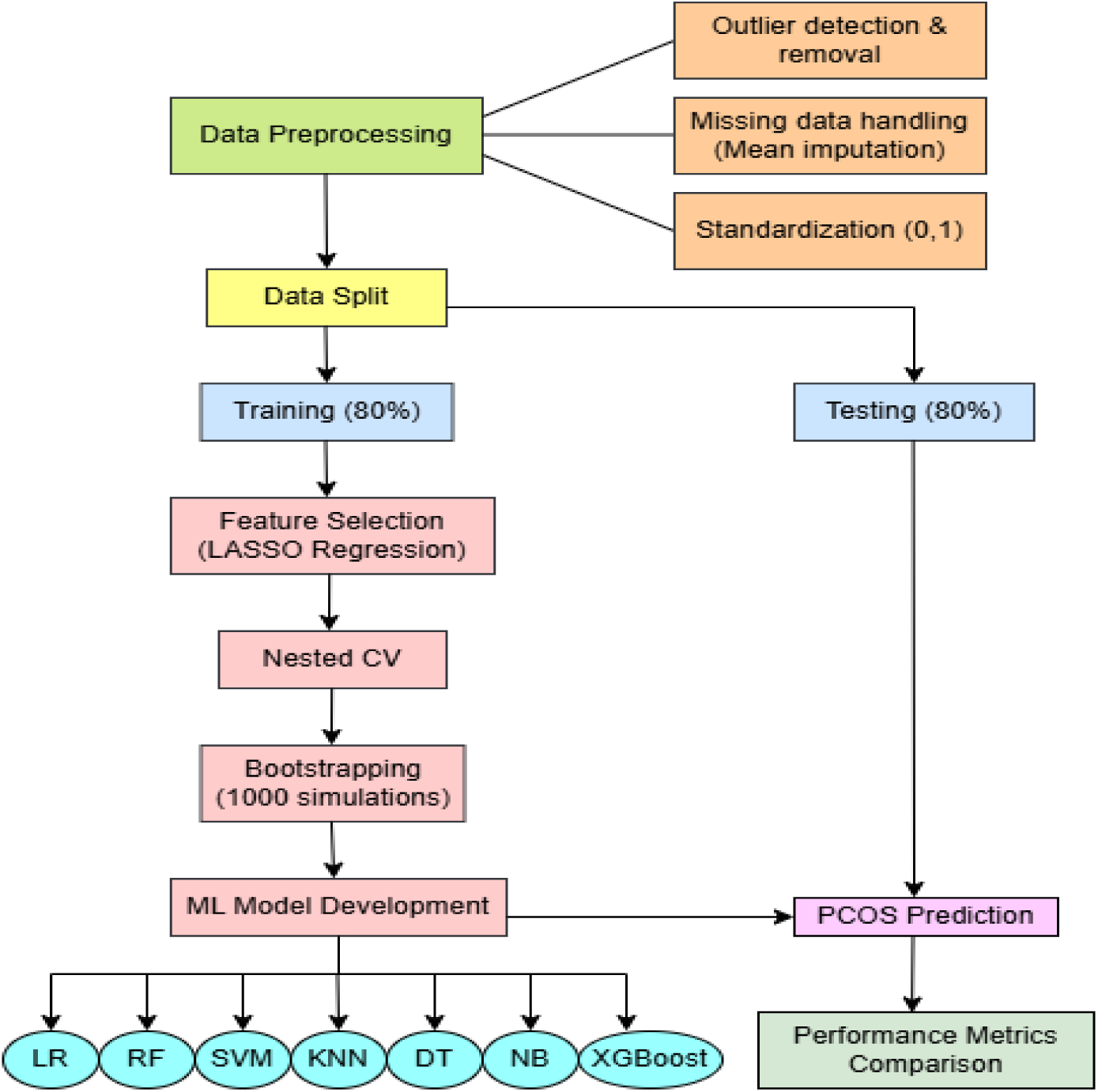
The workflow of the study.

### 2.8. Pre-processing and Feature Selection

Pre-processing was one of the important step that included identifying and eliminating outliers, handling missing values, and scaling data using standardization. In this study, missing values were addressed using mean imputation and after imputation, a stratified split was used to separate the dataset into training and testing subsets. Moreover, to improve comparability among predictors, all variables were standardized using scaling (zero mean and unit variance). Feature selection was performed using the least absolute shrinkage and selection operator (LASSO) regression. This process improved model interpretability and reduced the likelihood of overfitting by removing weak or irrelevant variables. A nested cross-validation (CV) was used to improve the reliability of model selection and performance estimation. For machine learning performance evaluation, a nested CV, which consists of two loops, is considered the gold standard since it prevents information leakage between the training and validation phases. Additionally, to increase the robustness of the estimates and decrease sampling variability, bootstrapping resampling was used to create multiple simulated datasets. After selecting features, a number of machine learning methods, such as Logistic Regression (LR), Random Forest (RF), Support Vector Machine (SVM), k-Nearest Neighbours (KNN), Decision Tree (DT), Naïve Bayes (NB), and Extreme Gradient Boosting (XGBoost), were trained to predict PCOS status. Feature importance was measured and visualized using performance metrics like as accuracy, sensitivity, specificity, the area under the receiver operating characteristic curve (ROC-AUC), and Cohen’s Kappa.

### 2.9. Nested Cross-Validation

To prevent overfitting and data leakage and ensure unbiased model evaluation and reliable feature selection, a nested cross-validation (CV) was implemented. This approach consists of two levels of cross-validation: an inner loop, which is used for hyperparameter tuning and an outer loop, which is used for model evaluation.

Outer loop: The dataset is divided into 5 folds. In each iteration, four folds were used for training, and the remaining fold was held out as the outer test set. Model performance was evaluated on this unseen fold, providing an unbiased estimate of generalizability.

Inner loop: Within each outer training set, a 5-fold inner CV was applied using Lasso CV to tune the regularization parameter (α) and identify the most important features. This step ensured that feature selection and hyperparameter optimization were conducted independently of the outer test set.

The average performance across all outer folds was calculated as:

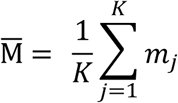

Where, M̄ is the final nested CV performance estimate, 𝑚_𝑗_ is the performance metric on the jth outer fold, and K is the total number of outer folds.

Feature stability: The final set of predictors was determined by a majority vote across the outer folds. The features that were consistently selected in the majority of outer iterations were kept for future model development.

This two-stage technique ensures that the outer test data is totally independent of the feature selection and hyperparameter optimization processes, eliminating information leakage and delivering an unbiased estimate of model generalizability [42].

### 2.10. LASSO(Least Absolute Shrinkage and Selection Operat or) with nested cross validation

LASSO regression was used to select important features by a penalized regression technique that increases the magnitude of regression coefficients by an L1 penalty. In order to eliminate non-informative predictors while keeping the most significant variables, this penalty reduces less significant coefficients towards zero.

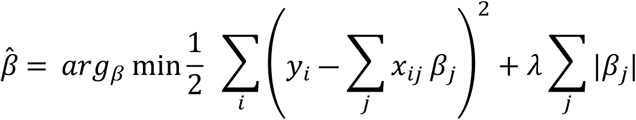

Where, y denotes the responses, X represents the matrix of predictors, β are the regression coefficients, and λ is the regularization parameter controlling the degree of shrinkage. Consequently, LASSO enhances the interpretability of the model and decreases the probability of overfitting, which is particularly crucial when dealing with several correlated predictors [43].

To find the best penalty parameter (𝜆), 5-fold cross-validation within the nested CV framework was used. At each iteration, models were fitted across a variety of λ values, and the cross-validated error was calculated. The optimal tuning parameter was determined by minimizing the mean cross-validated error with λ.

The study used LASSO regression within a nested cross-validation framework to improve the stability and dependability of variable selection, consistent with recommendations to combine penalized regression with resampling techniques [44]. Following the process, the final group of predictors was utilized for developing machine-learning models for PCOS prediction.

### 2.11. Bootstrapping

A bootstrapping method was used on the training data to ensure the models’ robustness and generalizability. Bootstrapping is a resampling method in which the original training set is randomly sampled with replacement to create multiple new datasets [45].

In this study, 1000 bootstrap simulations were performed. A bootstrapped dataset was used to train the machine learning models for each iteration, and a fixed independent test set was used for evaluation. This iterative resampling approach enabled to account for sampling variability and generate more accurate estimates of performance metrics [46]. The aggregated findings from all bootstrap iterations were summarized using mean values of evaluation metrics, leading to an adequate assessment of model generalizability. Deshmukh H et al. (2019) also used similar approach for the diagnosis and risk stratification of polycystic ovary syndrome (PCOS) [47].

### 2.12. Machine Learning Models

Table 2 presents the types of all machine-learning algorithms used in this study.

**Table 2.** Different machine learning algorithms with types.

|  | Algorithms |
| --- | --- |
| Types |  |
| Classical | Logistic regression (LR), Naive Bayes (NB),<br>Decision Tree (DT) |
| kernel-based | Support vector machine (SVM) |
| Instance-based | K-Nearest Neighbors (KNN) |
| Ensemble | Random forest (RF), extreme gradient boosting<br>(XGB) |

#### 2.12.1. Logistic Regression (LR)

Logistic Regression is a linear classification model that predicts the probability of a binary outcome by applying the logistic (sigmoid) function to a linear combination of input features. The regression function is:

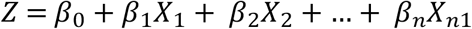

Where, 𝑍 is the predicted log-odds, 𝛽_0_is the intercept, and 𝛽_1_,…,𝛽_𝑛_ are the feature coefficients.

The probability of class membership was calculated using:

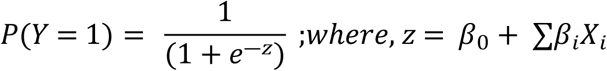

This transformed linear regression output into probabilities, making it suitable for binary classification [48].

#### 2.12.2. Support Vector Machine (SVM)

The Support Vector Machine (SVM) is a potent supervised learning technique that divides data points of several classes using the most optimal hyperplane. The primary goal is to optimize the margin, or the separation between the nearest data points from each class and the separating hyperplane. Better generalization is typically indicated by a larger margin. SVM can handle both linear and nonlinearly separable data by implicitly mapping inputs into higher-dimensional spaces using kernel functions [49]. In the linear form of SVM, it is defined as:

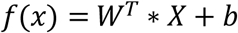

Where, X is the feature vector, W is the weight vector perpendicular to the hyper plane, and b is the bias term.

#### 2.12.3. Naive Bayes (NB)

Based on Bayes’ theorem, the Naïve Bayes is a probabilistic classifier and assumes conditional independence between features given the class. The equation for the Naive Bayes classifier is defined as:

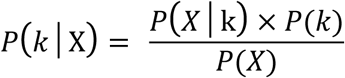

Where, 𝑃(𝑘│X) is the posterior probability of class k given the feature vector X, 𝑃(𝑋│k) is the likelihood, 𝑃(𝑘) is the prior, and 𝑃(𝑋) is the marginal probability.

Naïve Bayes is computationally efficient, utilizes minimal training data, and performs well even when the independence assumption fails to satisfy, making it helpful for prediction tasks where features are not necessarily highly correlated [50].

#### 2.12.4. K-Nearest Neighbors (KNN)

K-Nearest Neighbors (KNN) is a straightforward instance-based learning technique that is non-parametric. Instead of creating an explicit model, it uses the majority class of the closest neighbors in the feature space to categorize new data points. Euclidean distance is the most commonly used distance metric, which is calculated as follows:

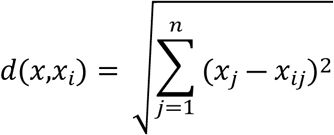

Where, x is the test instance, 𝑥_𝑖_ is a training instance, and *n* is the number of features.

KNN is a lazy learning process as it has no explicit training phase and it works well when decision boundary is irregular. However, it can be computationally expensive on large datasets [51].

#### 2.12.5. Decision Tree (DT)

A supervised learning approach called a decision tree creates a tree-like structure by iteratively dividing the data into subsets according to feature values. Every internal node stands for a feature test, every branch indicates the test’s result, and every leaf node represents a class label. To assess the quality of a split, DTs commonly use measures such as Gini Impurity. The Gini Impurity for a node is evaluated using the following formula:

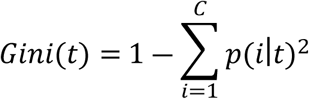

Where, 𝑡 is a node, 𝐶 is the number of classes, and 𝑝(𝑖∣𝑡) is the proportion of class i at node t. The Gini Impurity quantifies the extent of class mixing within a node; a node is considered purer if its Gini score is lower. However, DT is easy to interpret and can captures non-linear relationships. Nevertheless, it is prone to overfitting sometimes [52].

#### 2.12.6. Random Forest (RF)

Random Forest is an ensemble learning technique, which creates several decision trees and aggregates their results to get predictions that are more reliable and accurate. Each tree is constructed using a random subset of training data and a random subset of features at each split to reduce over-fitting and improve generalization [53]. Typically, the final prediction is decided by a majority vote among the trees. The final prediction is determined by:

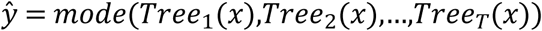

Where, 𝑇𝑟𝑒𝑒_𝑖_(𝑥) is the prediction from the 𝑖^𝑡ℎ^ decision tree, and 𝑇 is the total number of trees in the forest.

Random Forest is highly effective because it reduces variance compared to a single decision tree.

#### 2.12.7. Extreme Gradient Boosting (XGBoost)

XGBoost is a sophisticated gradient boosting method that sequentially constructs an ensemble of weak learners, usually decision trees. The final forecast is derived by combining the results of each new tree, which corrects the errors of the prior ensemble. The core of XGBoost lies in reducing a regularized objective function that includes both the training loss and a penalty term to control model complexity:

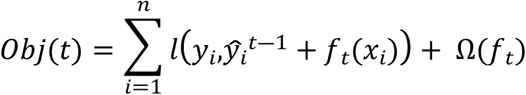

Where, 𝑙 is a differentiable convex loss function measuring the difference between prediction 𝑦_𝑖_^𝑡―1^ and the actual label 𝑦_𝑖_, 𝑓_𝑡_ represents the decision tree added at iteration 𝑡, and Ω(f) is the regularization term that penalizes tree complexity of 𝑓_𝑡_.

XGBoost uses column (feature) subsampling to increase variety, provides handling of missing values, and integrates L1 (Lasso) and L2 (Ridge) regularization (Ω (f)) to minimize overfitting [54].

#### 2.13. Evaluation Metrics

The predictive performance of the machine learning models was evaluated using a variety of standard classification measures to ensure a comprehensive evaluation. These included accuracy, sensitivity (recall), specificity, Cohen’s kappa coefficient, Receiver Operating Characteristic (ROC) Curve, and the area under the curve (AUC). All metrics were assessed using the confusion matrix, which consisted of True Positives (TP), False Positives (FP), True Negatives (TN), and False Negatives (FN).

#### 2.13.1. Accuracy

Accuracy is the overall proportion of correctly classified cases, calculated as the ratio of true positives and true negatives to the total number of cases [55].

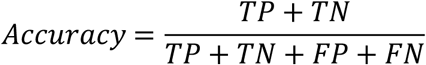

A high accuracy score indicates that the model can correctly predict both classes.

#### 2.13.2. Sensitivity

Sensitivity, also known as recall or the true positive rate defined as the proportion of true positives out of all actual positive cases [56].

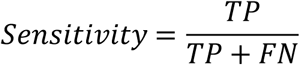

Sensitivity determines how well the model can detect PCOS individuals. A higher sensitivity means that the model rarely misses actual PCOS cases.

#### 2.13.3. Specificity

Specificity, also known as true negative rate defined as the proportion of true negatives out of all actual negative cases [57].

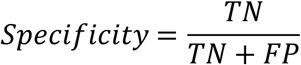

Specificity measures the model’s capacity to accurately classify individuals without PCOS.

#### 2.13.4. Cohen’s Kappa

Cohen’s Kappa is a chance-corrected measure of the agreement between predicted and observed classifications. Unlike accuracy, it considers the possibility of agreement appearing by chance [58].

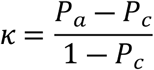

Where, 𝑃_𝑎_ is the observed agreement, and 𝑃_𝑐_ is the expected agreement by chance.

#### 2.13.5. ROC Curve and AUC

An ROC curve is a graphical representation that plots the true positive rate (sensitivity) against the false positive rate (1 − specificity) across a range of classification thresholds [59].

The Area Under the ROC Curve (AUC) was used as a summary measure of discriminative capacity. AUC values range from 0.5 to 1.0, where AUC = 0.5 indicating no discriminative ability (equivalent to random guessing) [60]. A greater AUC suggests that the model performs better at ranking PCOS patients higher than non-PCOS patients over a range of thresholds.

## 3.0 Results

Table 3 depicts Socio-demographic characteristics among reproductive-aged women in Bangladesh. All participants were reproductive-aged women, aged between 15 and 45. The participants in the study had an average age of 24.05 years (SD = 3.071), an average height of 166.10 inches (SD = 14.552), and a mean weight of 63.42 kg (SD = 20.108). The majority of the participants were single (56.1%) and came from rural settings (53.3%). Among the respondents, most had completed a graduation degree (62.7%). The comforting aspect was that most participants confirmed they were not affected by any disease (62.6%) and had no physical disability (89.2%). Moreover, the majority of them had no smoking or drinking habits (83.4%). As a result, the average BMI was determined to be 22.69 (5.434) among the respondents.

**Table 3.**
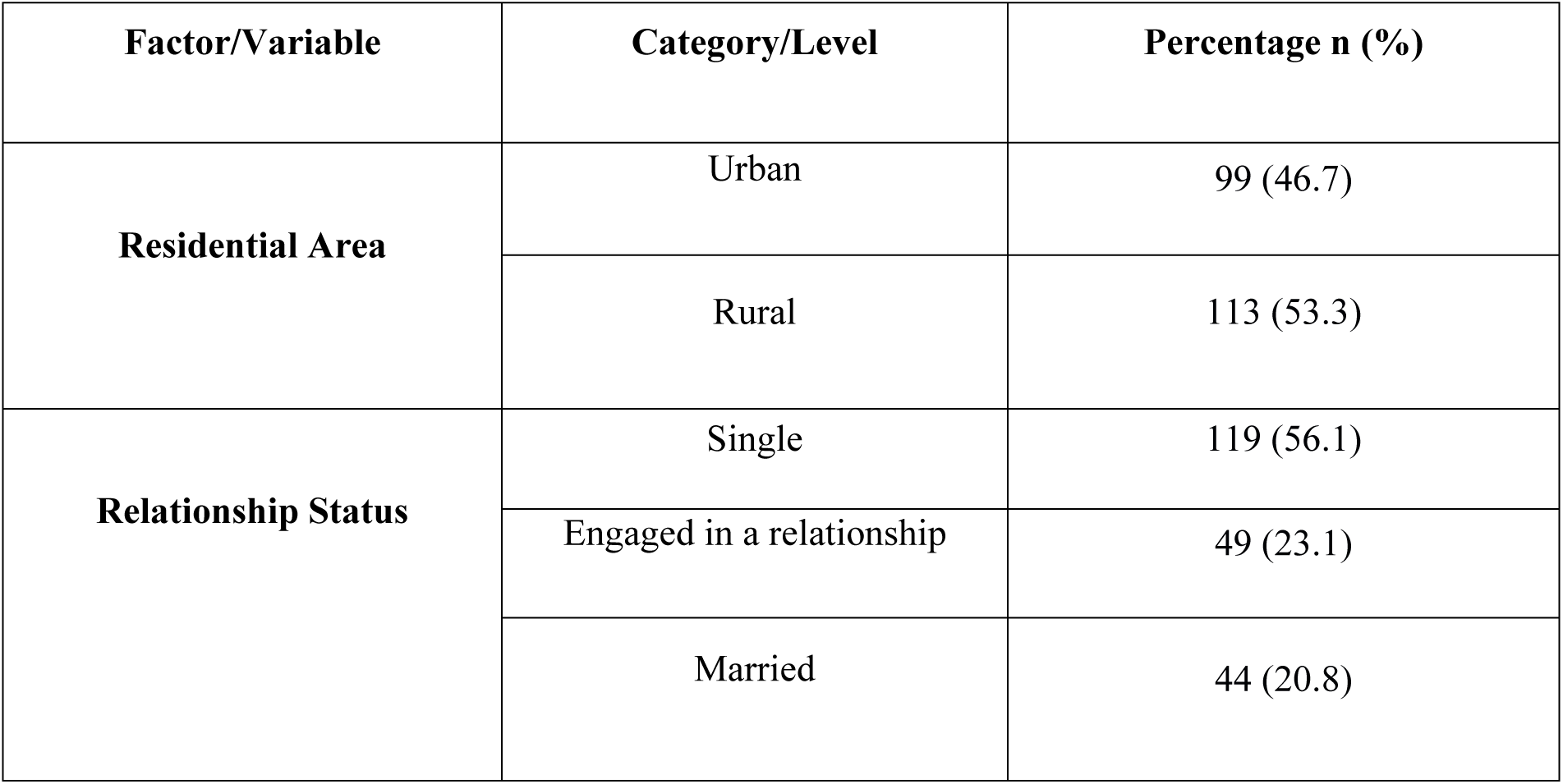
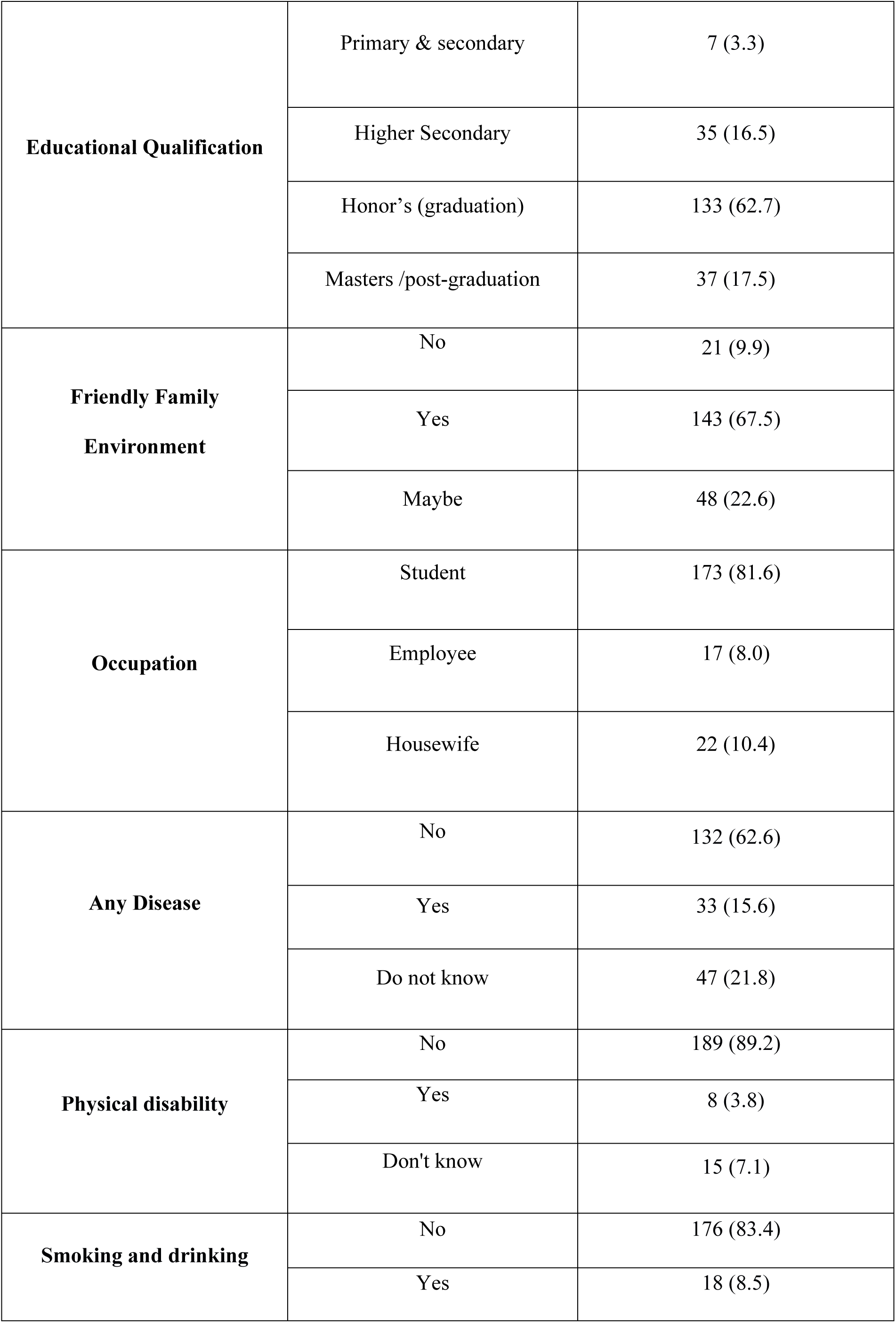
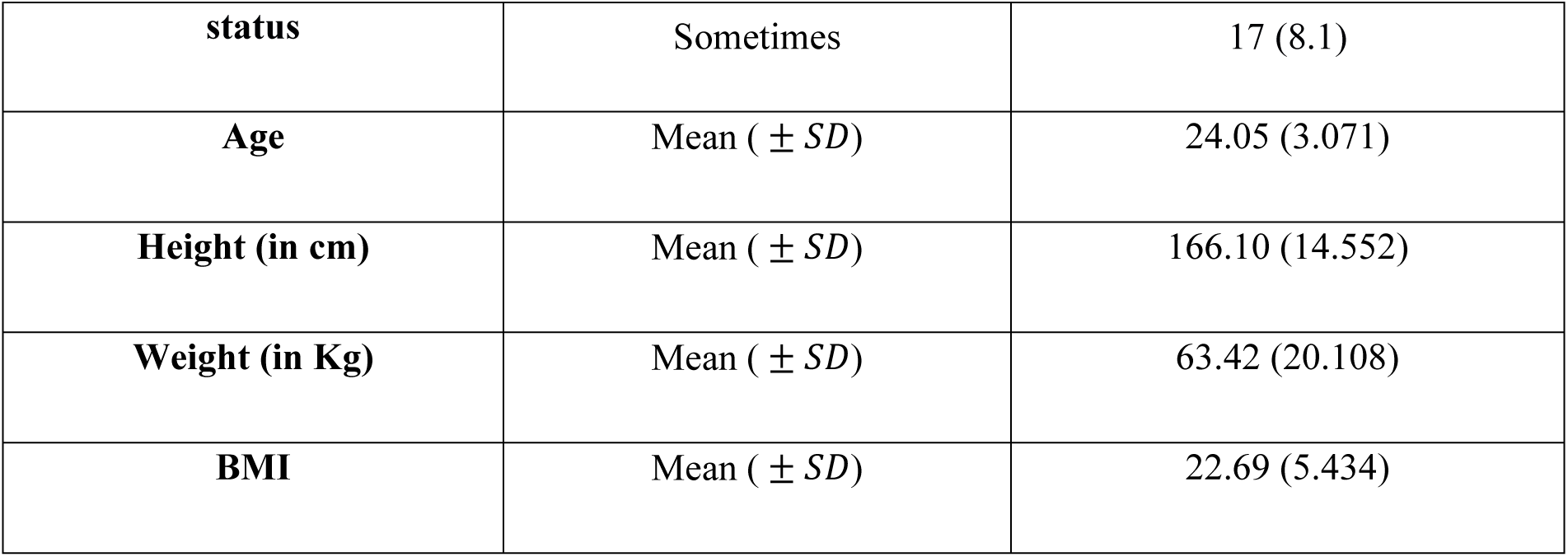
Socio-demographic characteristics of the Study Population in Bangladesh.

Table 4 illustrates PCOS and psychological factors related characteristics among reproductive-aged women in Bangladesh. The study demonstrated that the respondents had an average level of psychological outcome, including depression (14.76, 5.841), anxiety (15.82, 7.383), stress (18.17, 8.401), and insomnia (14.50, 5.203). Based on the findings of the Rotterdam criteria, an elevated level of irregular menstruation (50.9%) and hyperandrogenism syndrome (64.6%) was seen among the majority of participants. Furthermore, 41% of respondents used an ultrasound test to determine that each ovary contained 12 or more small follicles (2–9 mm in diameter).

**Table 4.**
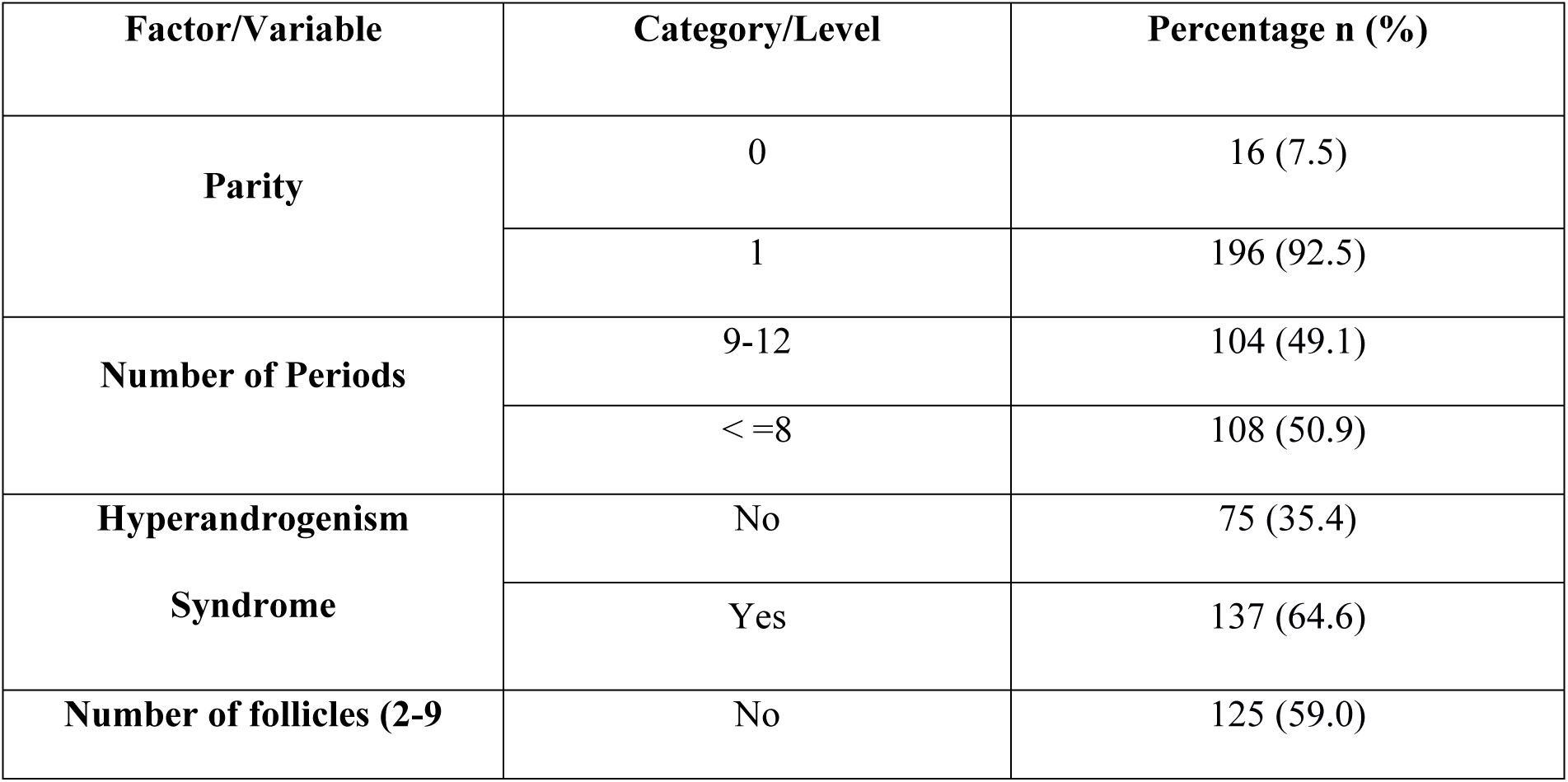
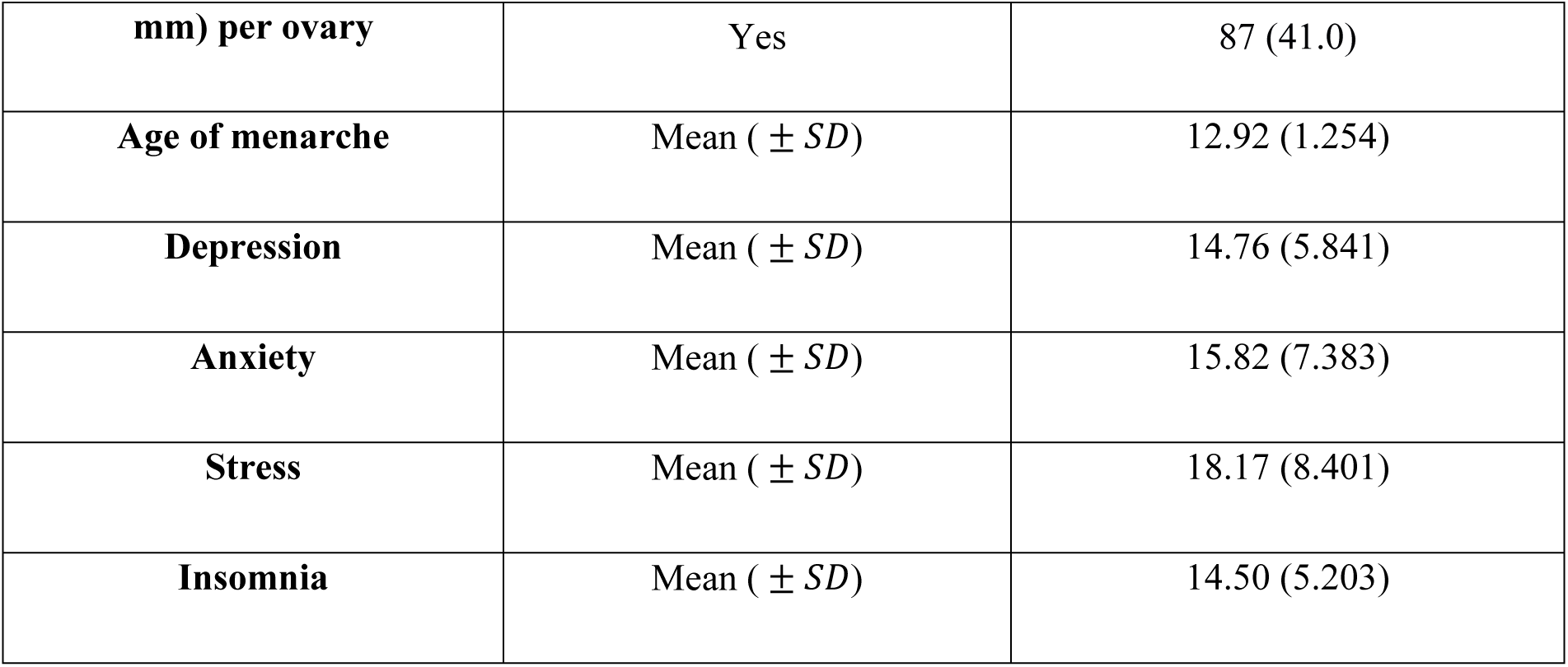
Health related Characteristics among reproductive-aged women in Bangladesh.

Figure 4 represents the prevalence of PCOS status among reproductive-aged women in Bangladesh. According to the Rotterdam criteria, around half of the participants were determined to be affected by polycystic ovarian syndrome (49.5%), and the remaining portion indicates those who did not have PCOS.

**Fig 4.**
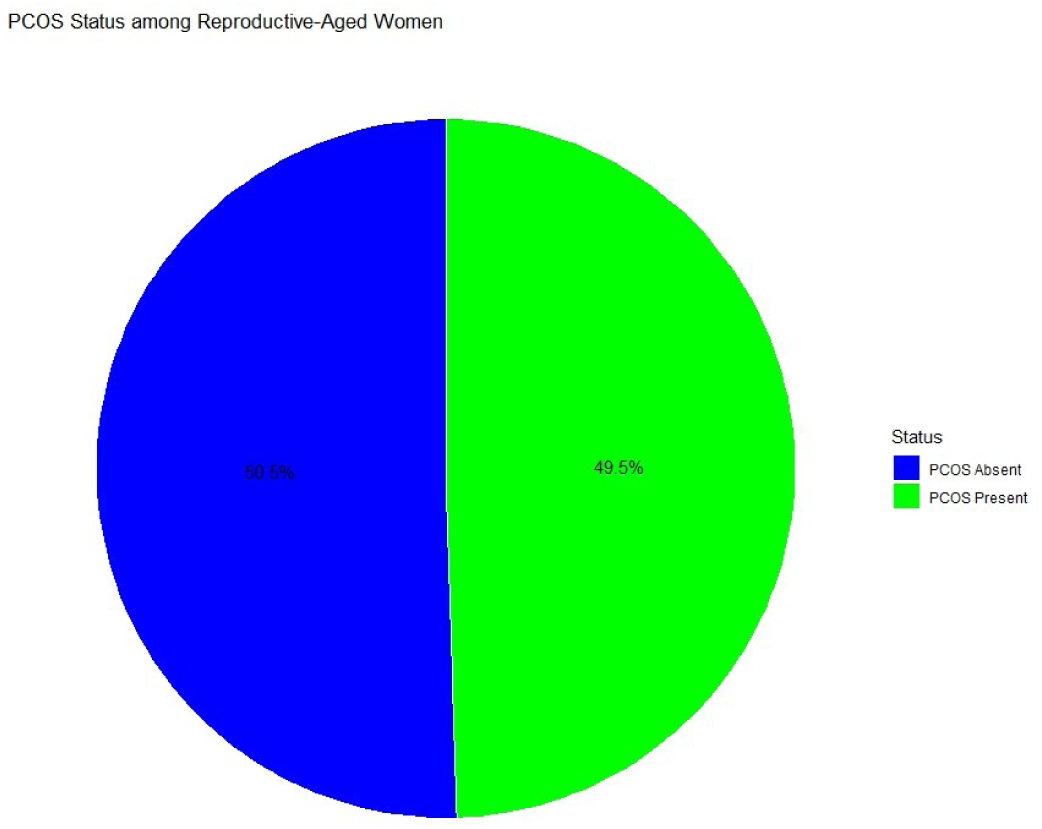
Prevalence of PCOS Status among Reproductive-Aged Women in Bangladesh.

Figure 5 displays the most essential features selected by Lasso regression. Initially, 21 features were used in the study. After conducting Lasso regression, eight features were chosen from 21, as they were the most influential for predicting PCOS by eliminating less essential features. Among the eight features, the number of periods (0.12), hyperandrogenism syndromes (0.15), and the number of follicles per ovary (0.22) had the highest coefficient of magnitude, indicating that these predictors were the most influential in predicting PCOS. Moreover, two psychological outcomes, including depression and stress, were relatively less significant contributors that had a lower impact on predicting PCOS compared to the first three features. Finally, the socio-demographic variables, like any other disease (0.03) and educational qualification (0.04), had the lowest coefficient of magnitude, indicating that those features had minimal influence on the model’s prediction.

**Fig 5.**
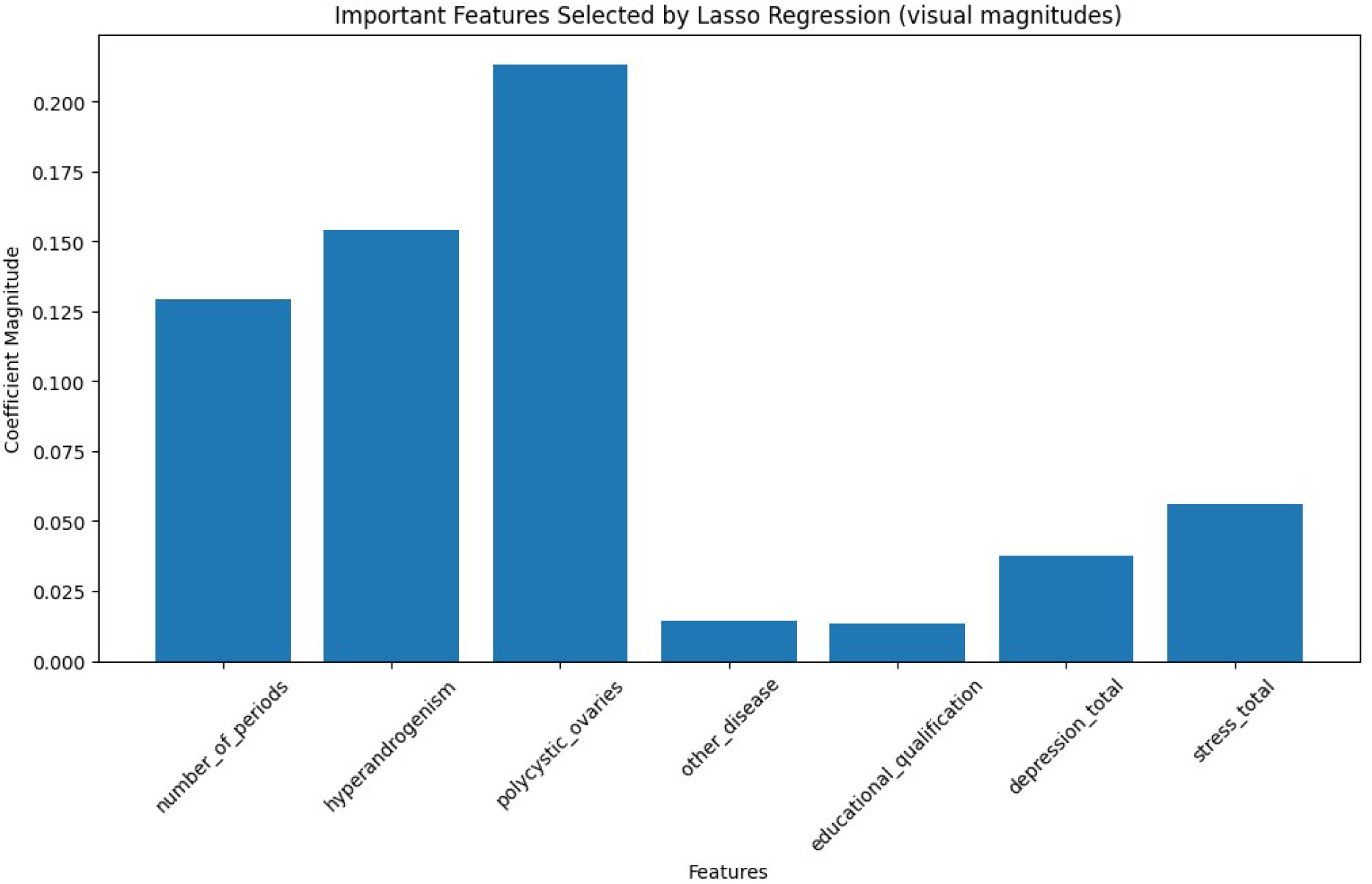
Important features selected by LASSO Regression.

Fig 6 depicts the Correlation Matrix Heatmap of study variables. The Correlation heatmap revealed that PCOS diagnosis status was positively and strongly linked with clinical parameters such as polycystic ovaries (0.82), hyperandrogenism (0.60), and number of periods (0.67). Furthermore, socio-demographic indicators such as occupational status (0.54) and relationship status (0.45) had a moderate correlation with PCOS diagnosis status. Furthermore, a weak moderate positive correlation was observed between PCOS diagnosis status, educational qualification (0.29), and residential area (0.32). Similarly, PCOS status had a very weak correlation with depression (0.06). However, a weak and negative association was found between PCOS diagnosis status and insomnia (-0.14). The data indicated that PCOS diagnosis was most strongly associated with clinical aspects (irregular periods, polycystic ovaries, hyperandrogenism), while psychological components had moderate to weaker associations.

**Fig 6.**
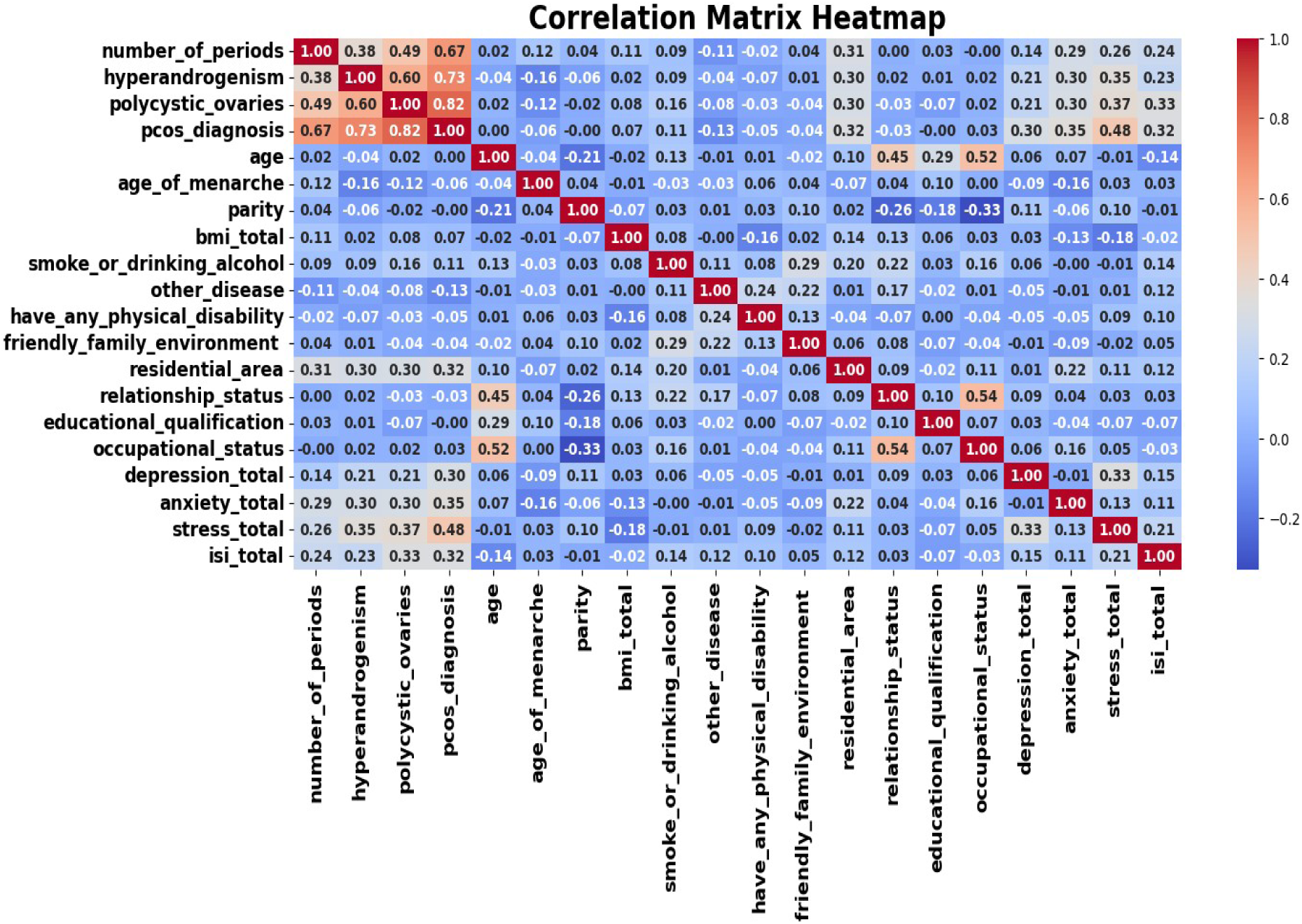
Correlation Matrix Heatmap of study variables.

Table 5 presents a comparative analysis of various machine learning models in predicting PCOS. The models were evaluated based on several key matrices, including accuracy, sensitivity, and specificity, kappa, and ROC-AUC value.

**Table 5.** Comparative performance of different machine learning models in predicting PCOS.

| **Models** | **Accuracy (%)** | **Sensitivity (%)** | **Specificity (%)** | **Kappa Score (%)** | **ROC-AUC (%)** |
| --- | --- | --- | --- | --- | --- |
| XG-Boost | 99.631 | 99.450 | 99.812 | 99.262 | 99.999 |
| RF | 99.264 | 98.550 | 99.978 | 98.528 | 99.994 |
| SVM | 98.975 | 98.084 | 99.865 | 97.950 | 99.999 |
| KNN | 97.631 | 96.865 | 98.397 | 95.262 | 99.321 |
| DT | 98.056 | 97.512 | 98.600 | 96.112 | 98.056 |
| LR | 98.254 | 96.587 | 99.921 | 96.509 | 99.846 |
| NB | 87.789 | 98.087 | 77.490 | 75.578 | 99.727 |

### Extreme Gradient Boosting (XG-Boost)

The XG-Boost model achieved an accuracy of 99.631%, indicating that it could classify 99.631% of instances accurately. Furthermore, the model demonstrated 99.450% sensitivity and 99.812% specificity, suggesting that it was capable of detecting 99.450% of positive and 99.812% of negative cases. Additionally, a kappa score of 99.262% was obtained in this model, indicating almost perfect agreement between the predicted and actual outcomes.

### Random Forest (RF)

The RF model successfully classified 99.264% of cases. Besides, this model accurately detected 98.550% of positive cases and 99.978% of negative cases, demonstrating that the model was more perfect in detecting the negative cases than positive cases. Moreover, an almost perfect agreement was found between actual and predicted outcomes.

### Support Vector Machine (SVM)

The SVM model was capable of accurately classifying 98.975% of instances. The model also showed better performance in detecting negative cases (98.084%) compared to positive cases (99.865%). Additionally, this model achieved almost perfect agreement (97.950%) between actual results and model predictions.

### K-Nearest Neighbors (KNN), Decision Tree (DT), and Logistic Regression (LR)

The KNN, DT, and LR algorithms achieved an almost consistent accuracy, indicating that the models were proficient in classifying accurately 97.631%, 98.056%, and 98.254% of cases, respectively. The three models also performed better in identifying negative cases (KNN = 96.865%, DT = 97.512%, LR = 96.587%) than in identifying positive cases (KNN = 98.397%, DT = 98.600%, LR = 99.921%). Moreover, an almost perfect agreement was also observed between actual findings and model predictions among these three models; however, the scores were lower than those of the previous three models, such as XG-Boost, RF, and SVM.

### Naive Bayes (NB)

The NB model could classify 87.789% of instances, which was relatively lower compared to other models. Additionally, this model exhibited higher sensitivity (98.087%) and lower specificity (77.490%), indicating that it had a greater ability to identify positive cases compared to negative cases. Additionally, the model achieved a substantial agreement (75.578%) between model predictions and actual results.

Fig 7 depicts the combined ROC plots for all the models. Among all the models, the XG-Boost algorithm achieved the highest ROC value, indicating a 99.999% greater ability to differentiate between PCOS-present and PCOS-absent classes. Furthermore, the remaining models achieved higher ROC values, indicating that RF, SVM, KNN, DT, LR, and NB had 99.994%, 99.999%, 99.321%, 98.056%, 99.846%, and 99.727% ability to differentiate between PCOS and non-PCOS groups, respectively. However, these models had a smaller ROC area under the curve (AUC) than the XG-Boost model.

**Fig 7.**
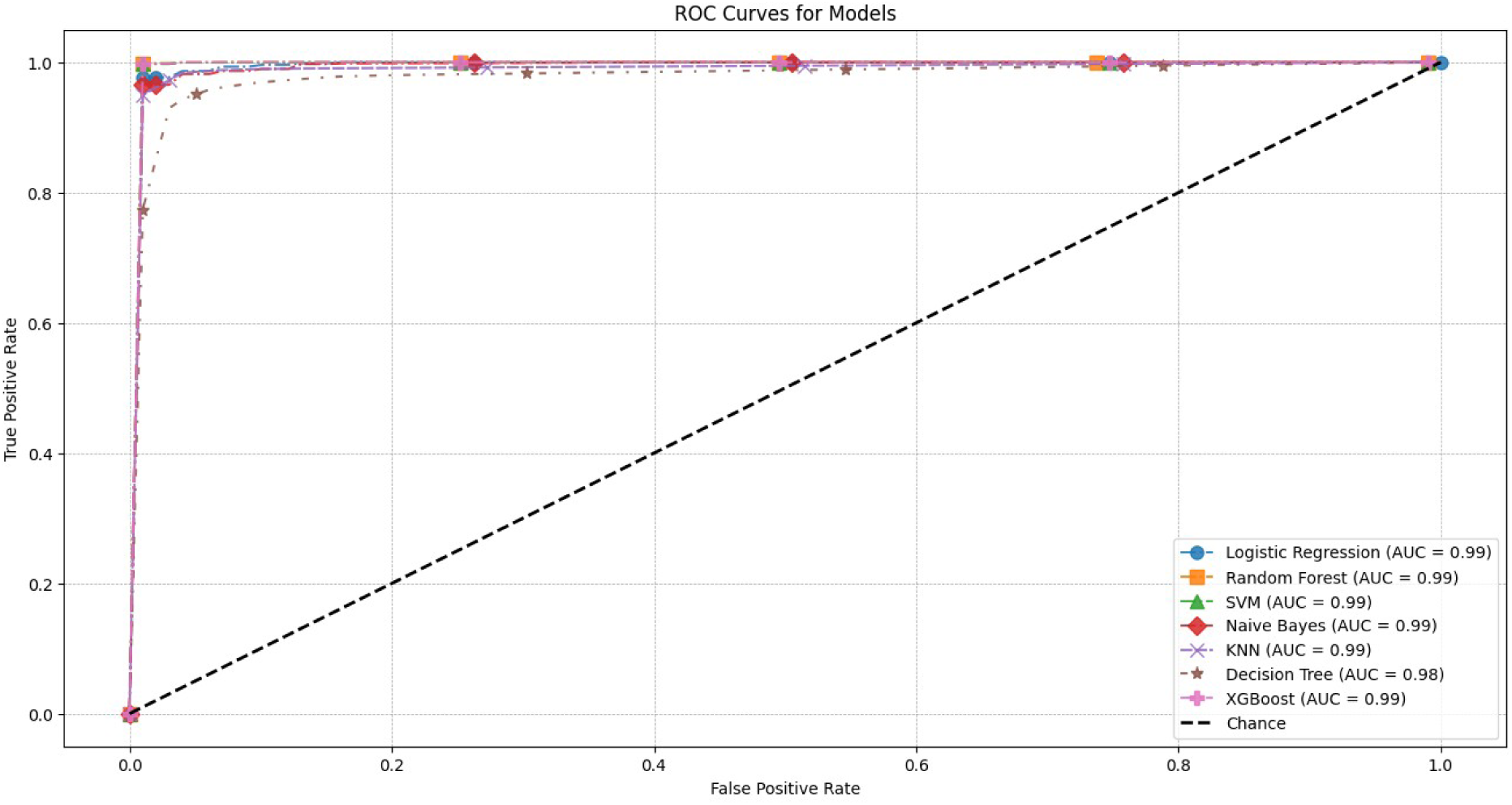
Combined Receiver Operating Characteristic Plots for all the Models.

Based on a comparison of the above models, XG-Boost was the best-fitting model, as indicated by the performance of the evaluation matrices. It outperformed the other models in terms of accuracy, kappa score, and ROC area under the curve values. Furthermore, due to its balanced nature, this model demonstrated the best ability to detect both PCOS and non-PCOS instances. As a result, the XG-Boost model was the best among all the standard models.

## 4.0 Discussion

This study utilized advanced machine learning (ML) methodologies aimed at developing a predictive model for polycystic ovary syndrome (PCOS) among reproductive-aged women in Bangladesh. The findings demonstrated excellence in diagnostic performance by integrating clinical and psychological dimensions related to PCOS. The outcomes of the study affirmed the contribution of advanced ML algorithms for disease prediction along with highlighting the socio-clinical determinants of PCOS that are relevant to health policy and clinical practice in low-resource settings.

According to the study, nearly half (49.5%) of the participants met the Rotterdam criteria for PCOS, reflecting a notable burden among this population. The observed prevalence for this study corresponds with numerous South Asian hospital-based investigations that applied the Rotterdam criteria and reported elevated prevalence rates of PCOS in both Bangladesh and India [61,62]. Compared to many international estimates, our observed prevalence was significantly higher, which underscores both diagnostic precision and the likelihood of increased predisposition among Bangladeshi women.

Clinical indicators such as irregular menstruation, hyperandrogenism and polycystic ovarian morphology in our models emerged as the strong predictive contributors. The findings provided consistency with the latest diagnostic framework, which validated the models clinical utility and consistency with the Rotterdam criteria and recent machine learning evidence [63,64]. Moreover, psychological features, particularly depression and stress, were sustained with minimal contribution, which validated a relatively weaker predictive role for PCOS. This finding aligned with a prior study which was conducted in China on women with PCOS [65]. Additionally, socio-demographic variables including educational qualification and the status of other disease were also attained but with a modest predictive capacity. This statement supported a previous study that found educational qualification and health status to be significant predictors of increased levels of PCOS [66]. This model provided evidence that suggested their potential utility as cost-effective features related to socio-psychological indicators, which might be used as a primary screening tool for identifying PCOS diagnoses in resource-limited settings like Bangladesh, where hormonal assays or transvaginal ultrasound tools were might be unavailable.

The incorporation of socio-demographic factors including occupational and relationship status which was moderately correlated to the risk of PCOS, highlighted the complex etiology of the syndrome. These associations were in accordance with emerging research that linked occupation and marital status to endocrine disturbances and reproductive dysfunction [66,67]. However, psychological variables like depression, anxiety, stress and insomnia exhibited a weaker but notable association with PCOS according to the findings of the study. These interrelations aligned with global evidence which demonstrated an association between PCOS and poor mental health outcomes [68,69]. The models used in this investigation adequately captured these clinical predictors along with additional socio-psychological dimensions, referring to a multidimensional approach.

The remarkable performance of extreme gradient boosting (XG-Boost) in this study reflected as the emerging consensus that outperformed traditional statistical and single-model ML approaches in clinical prognostication tasks [70]. A prior study reported that among diverse predictor variables, XGBoost achieved higher accuracy by modeling complex, non-linear relationships and interactions while maintaining a balanced trade-off between sensitivity and specificity [71]. It was consistent with the current findings which illustrated that XGBoost excels beyond simpler models in differentiating between PCOS positive and negative cases, achieving an accuracy of 99.631%, Cohen’s Kappa score of 99.262%, ROC AUC of 99.999%, sensitivity of 99.450%, and specificity of 99.812%. Precise identification of affected individuals reduces both false positives which minimize unnecessary anxiety and healthcare expenditures and false negatives for enabling timely clinical interventions. Moreover, classifiers such as Random Forest (RF) and Support Vector Machine (SVM) also attained impressive accuracies of 99.264% and 98.975% respectively, validating the effectiveness of ensemble and kernel methods in PCOS prediction. However, the comparatively lower performance of these two models validated the superiority of XGBoost in diagnostic ML modeling [72]. These findings pointed to the implementations of machine learning (ML) models which is capable of accommodating complex and intercorrelated clinical variables in the development of diagnostic systems.

This study used extensive clinico-psychological data to construct an interpretable machine learning framework for predicting PCOS. The intensive implementation of nested cross-validation, which is the gold standard for evaluating ML models, and 1000 bootstrapping simulations, ensured that our high-performance measures are robust to sampling fluctuation and internal data leakage. In addition, the model is only used as a non-serological and non-invasive information means, making it a low-cost and easily deployable screening tool capable of use in primary care settings in which there may be limited resources. The inclusion of both clinical parameters and validated psychological measures, such as DASS 21 and ISI 7, adds to the capability of capturing the multifactorial determinants of PCOS and further enhances its clinical relevance. Moreover, integration of the information of a cohort of Bangladeshi reproductive women fills a critical gap in evidence in LMIC settings and for working towards the development of a context-appropriate prediction model for real-world use.

Although the present study employed a rigorous analytic framework and demonstrated exceptional internal validity, several limitations must be acknowledged to contextualize the findings realistically. The primary limitation of this study was the small number of observations, which probably reduced the prediction capacity of all the trained models that are intended to operate with a higher number of events. However, the approach used for separating the training and validation sets may worsen the sample size issue. This may be addressed by using nested cross-validations, which keep the validation set totally blind for the model. The cross-sectional design of the study and limited regional dataset may introduce bias and restrict generalizability. Key predictors such as menstrual irregularity, hyperandrogenic signs, and ovarian follicle count are integral to the diagnostic criteria for PCOS, which may have inadvertently inflated model performance due to partial circularity. Despite the strict hyperparameter tuning and nested cross validation practice adopted to control the risk involved, the remarkable performance of tree-based ensemble algorithms such as XGBoost on a small dataset brings the risk of overfitting through memorization of data intricacies that cannot be conclusively ruled out without external validation. While near-perfect internal metrics were produced in this model, it is necessary to recognize that the high level of performance might be partially attributed to the high predictive capability of the PCOS diagnostic criteria that were used as features, which qualifies caution in the interpretation and extrapolation of the results. Finally, the absence of external validation and calibration assessment means that the real-world applicability of the model cannot yet be established.

## 5.0 Conclusion and Recommendations

This study enhances our understanding of how data-driven intelligence can support traditional diagnostic methods for polycystic ovary syndrome (PCOS) in low- and middle-income settings. By integrating demographic, clinical, and psychological factors into a cohesive analytical framework, it demonstrates that modern ensemble learning algorithms, particularly extreme gradient boosting can achieve remarkable predictive accuracy in identifying PCOS among women of reproductive age. Notably, feature selection using LASSO emphasized the clinical relevance of the prediction process, identifying menstrual irregularity, hyperandrogenic manifestations, and ovarian morphology as the primary indicators, while also revealing the significant yet secondary roles of psychological distress and socio-demographic context.

Prospective validation within routine gynecological and endocrinological care will help assess usability, cost-effectiveness, and acceptability among healthcare professionals. Simultaneously, policymakers and health planners in Bangladesh and similar regions could utilize this predictive approach to develop targeted awareness programmes, improve diagnostic protocols, and optimize resource allocation for women’s reproductive health. Further research should focus on determining the XGBoost model’s clinical significance, which will be followed by multi-center and external prospective validation of the model in various ethnic and geographic cohorts. Once these steps are achieved, machine learning–based frameworks could significantly transform the diagnostic landscape for PCOS and contribute to the broader agenda of data-informed public health innovation in resource-limited settings.

## Funding

The authors received no specific funding for this work.

## Declaration of competing interest

The authors ensure no conflicts of interest or personal relationships that could have appeared to influence the work reported in this paper.

## Data Availability Statement

All data are in the manuscript and/or supporting information files.

---

**SI Fig.**
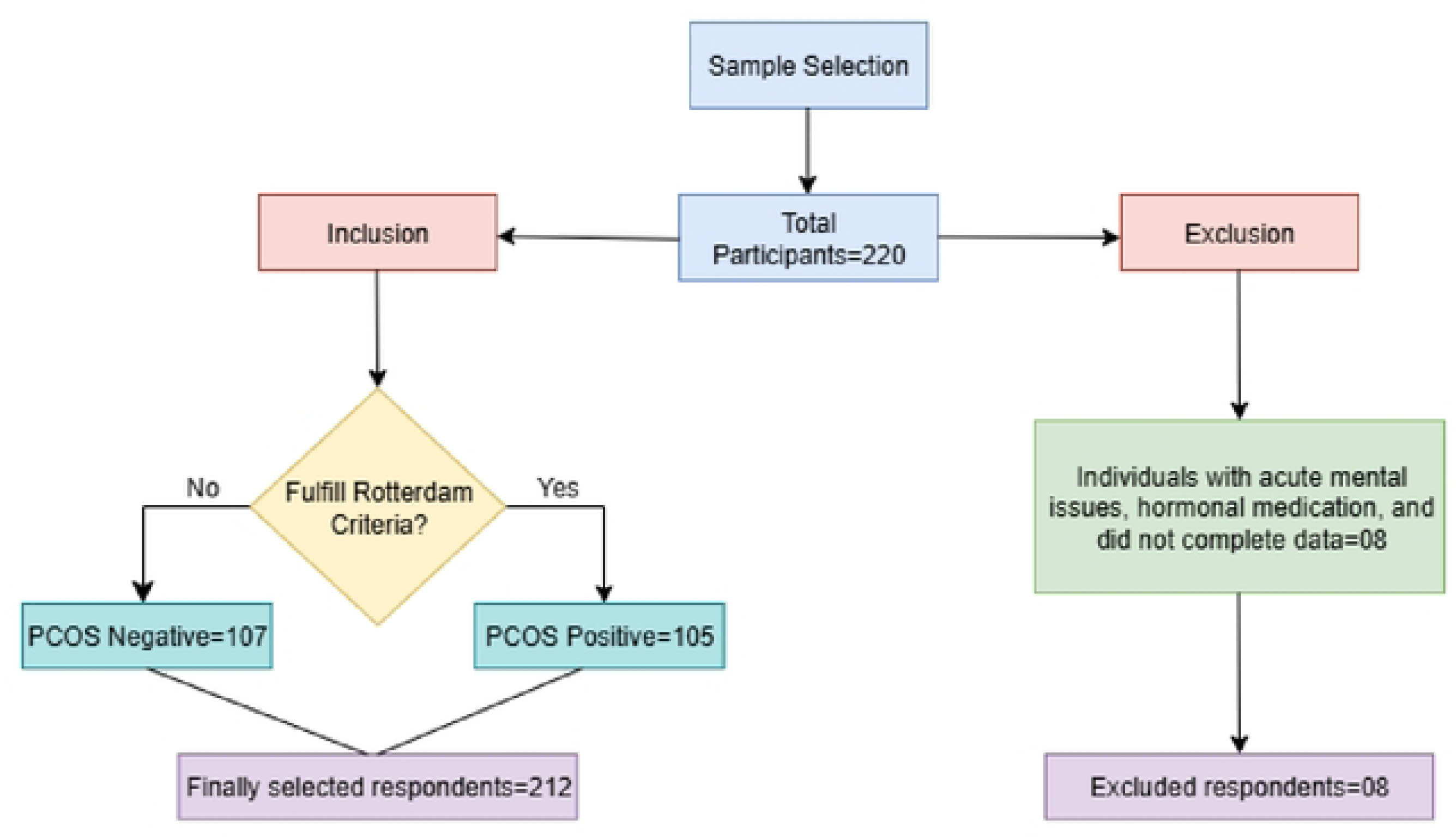
Sampling Flow chart with study inclusion and exclusion criteria.

**S2 Fig.**
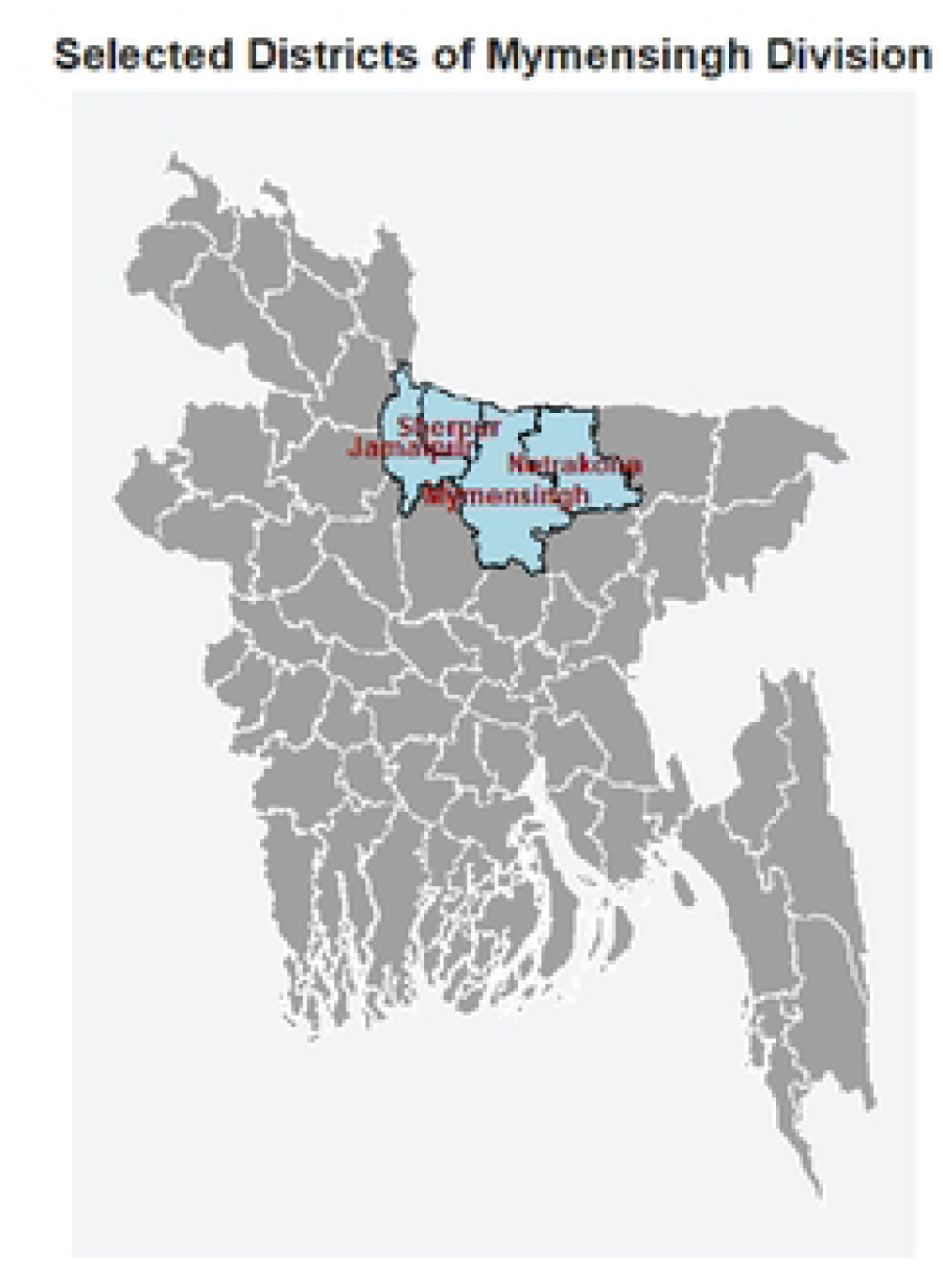
Map of the study area.

**S3 Fig.**
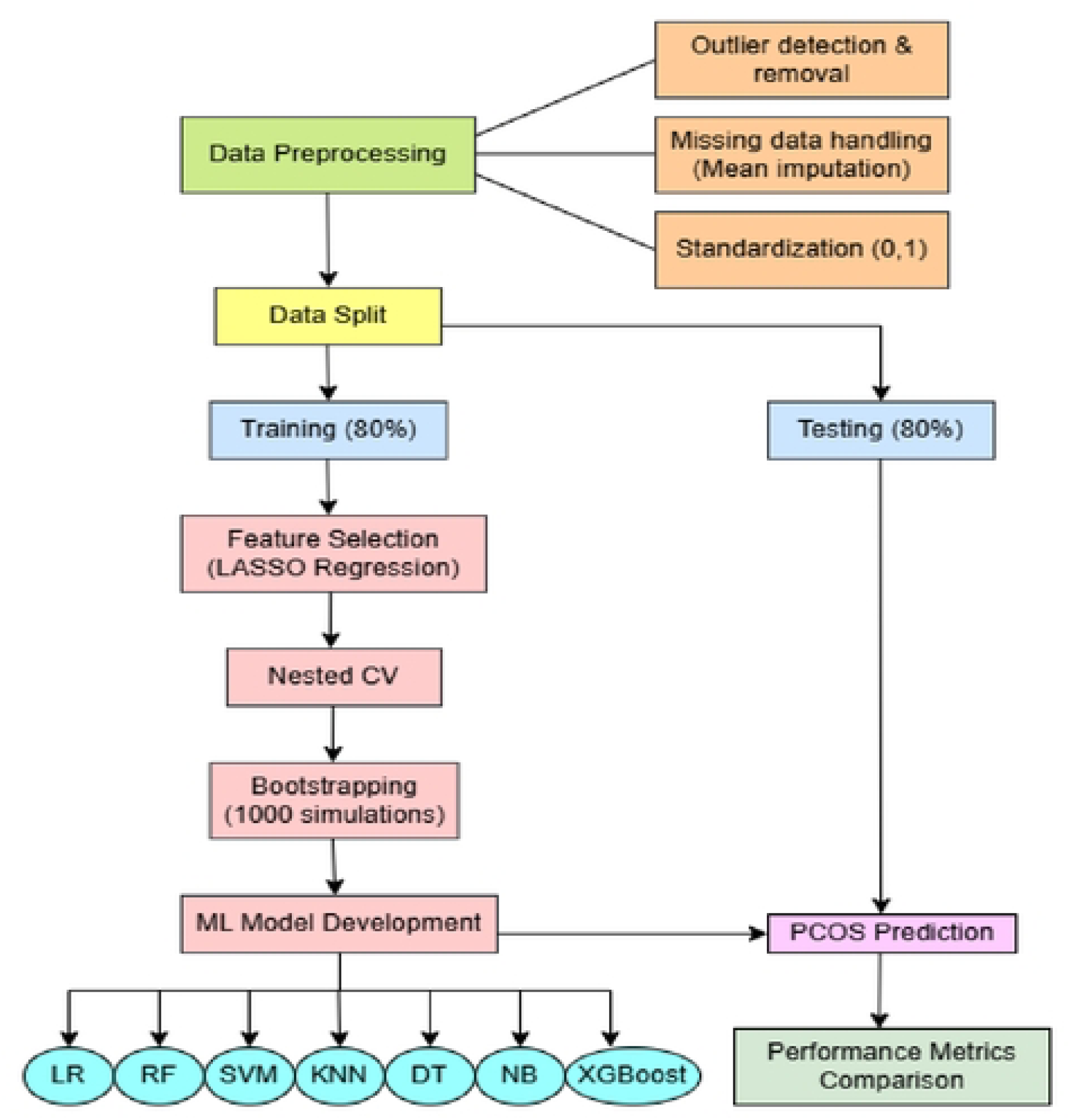
The workflow of the study.

**S4 Fig.**
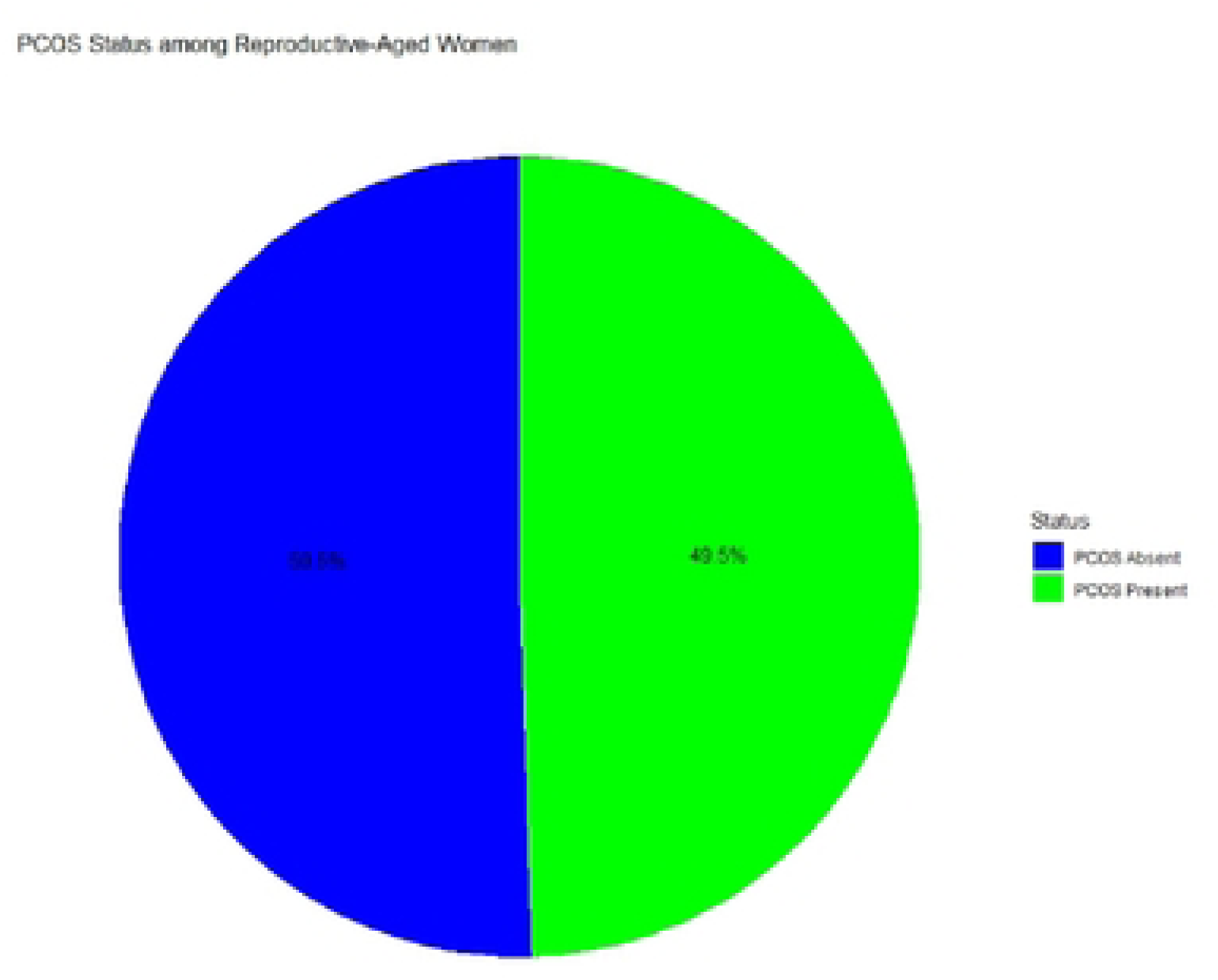
Prevalence of PCOS Status among Reproductive-Aged Women in Bangladesh.

**S5 Fig.**
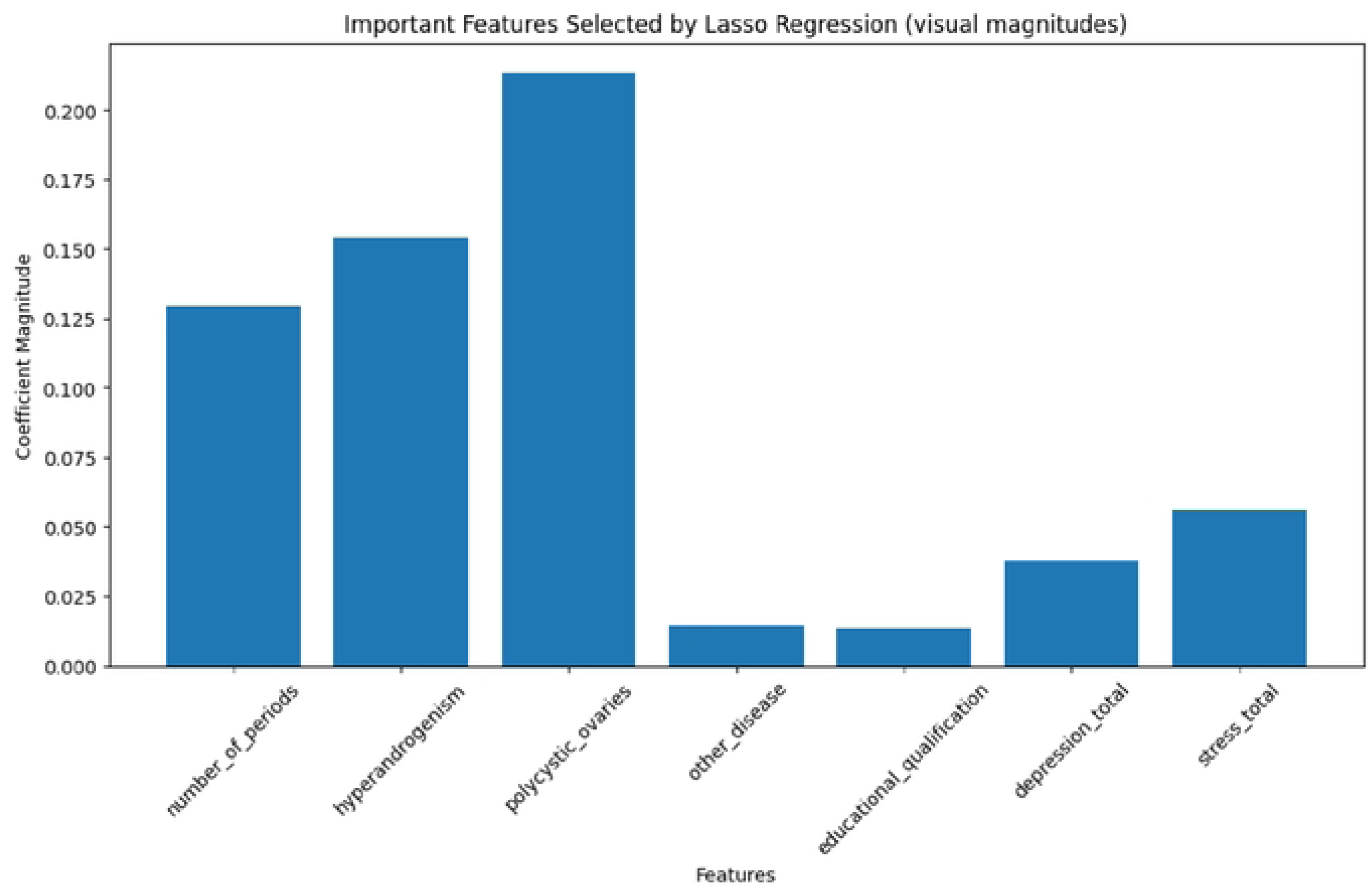
Important features selected by LASSO Regression.

**S6 Fig.**
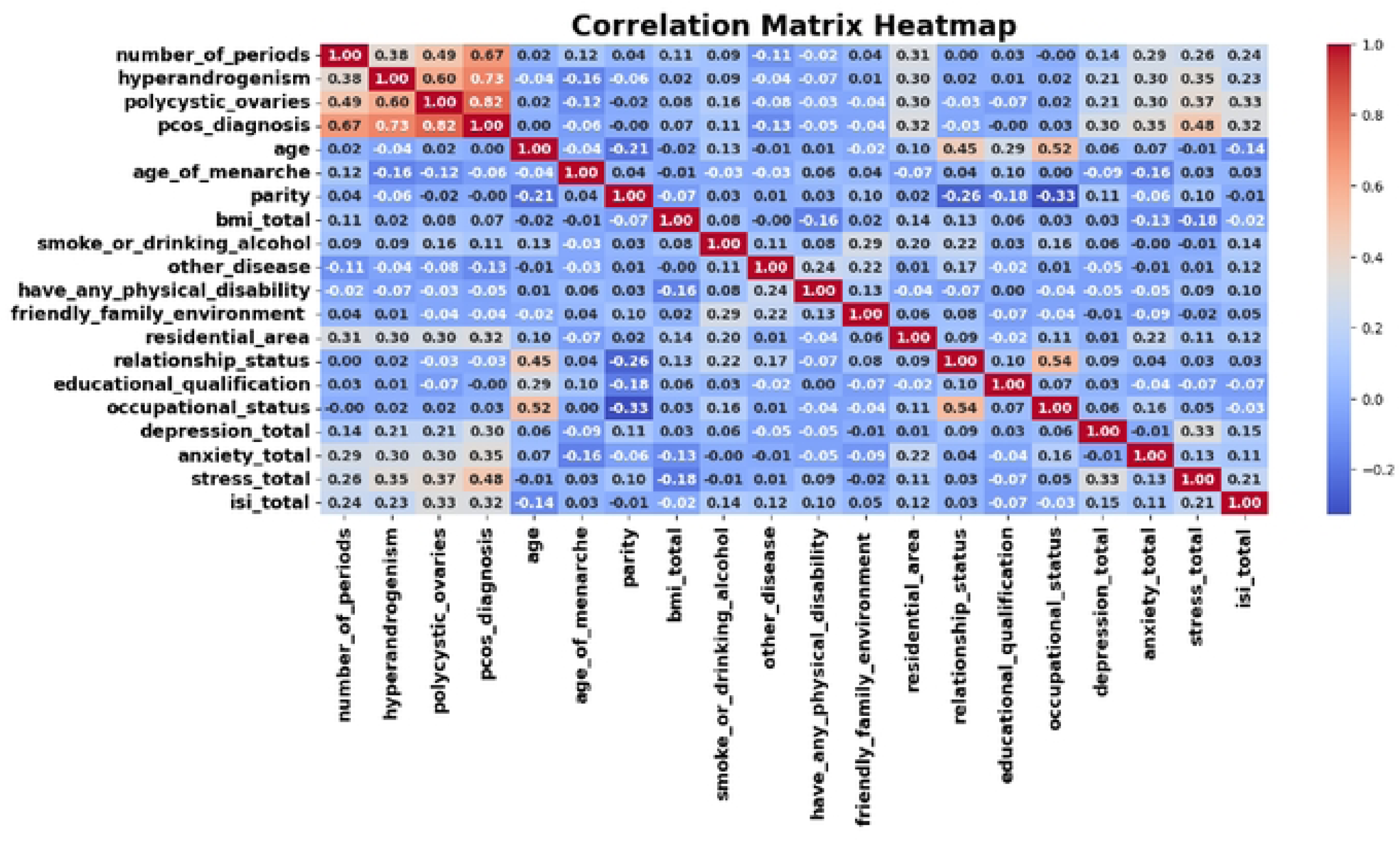
Correlation Matrix Heatmap of study variables.

**S7 Fig.**
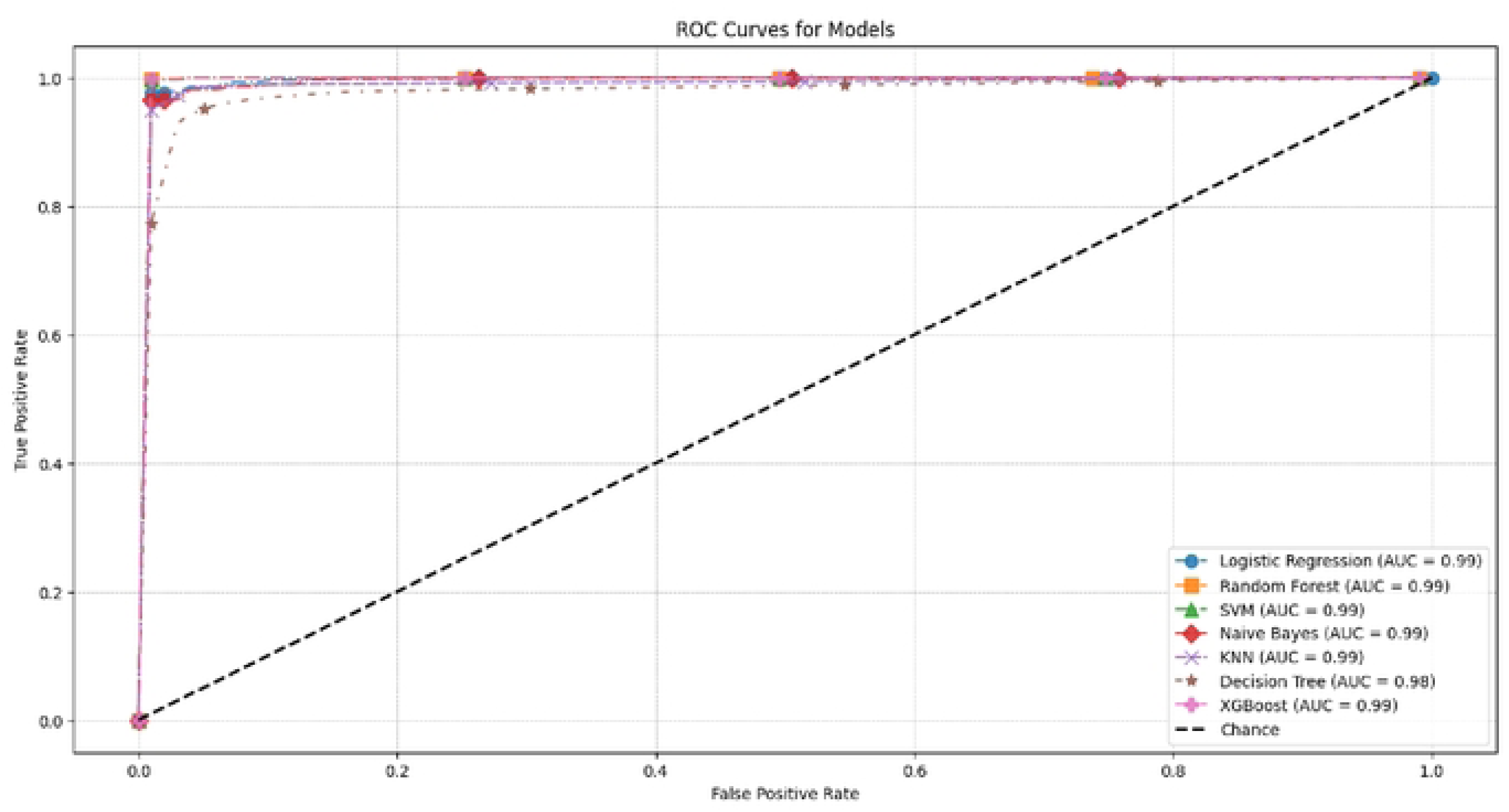
Combined Receiver Operating Characteristic Plots for all the Models.

## Notes

### Competing Interest Statement

The authors have declared no competing interest.

### Funding Statement

The author(s) received no specific funding for this work.

### Author Declarations

Ethical approval for this study was granted by the Research Ethics Committee of the Public Health Foundation Bangladesh (PHFBD) in Dhaka. The study adhered to the ethical principles outlined in the Declaration of Helsinki. Written informed consent was obtained from all participants prior to data collection. The research protocol was approved under reference number PHFBD-ERC-NFP-E-18/2025. At the beginning of data collection, participants received a thorough explanation of the study's objectives, aims, methods, affiliations, benefits, and hazards at the planned face to face interview session. Afterward, the respondents agreed to take part in the study, and the data collectors promised that their answers would be kept private and not shared. With their consent, data was subsequently gathered from them for the research purpose.

